# Apolipoprotein E genotype and MRI-detected brain alterations pertaining to neurodegeneration: A systematic review

**DOI:** 10.1101/2021.01.20.21250005

**Authors:** Albert Dayor Piersson, Mazlyfarina Mohamad, Subapriya Suppiah, Nor Fadilah Rajab

## Abstract

**Introduction:** The effect of apolipoprotein E (APOE) genotype, particularly APOE ε4, the main genetic risk factor for late-onset Alzheimer’s disease (LOAD), has been widely explored in neuroimaging studies pertaining to older adults. The goal of this systematic review was to review the literature on the relationship between carriage of the APOE ε4 allele and grey matter (GM) changes across various age groups and its influence on neurodegeneration as evidenced by structural magnetic resonance imaging (MRI).

**Methods:** A search of the electronic databases Pubmed, Scopus, Ovid and Cochrane was carried out till March 2020. Only studies published in English were included. Risk of bias of each study was assessed using the modified Newcastle-Ottawa Scale.

**Results:** A total of 115 articles met the inclusion criteria. Methodological quality varied from poor to good. There is moderate evidence of reduced GM volume in the middle frontal gyrus, precuneus, hippocampus, hippocampal subfields, amygdala, parahippocampal gyrus, middle temporal lobe, whole temporal lobe, temporal pole, and posterior cingulate cortex in APOE ε4 carriers.

**Conclusion:** The present data supports the utility of the hippocampal GM volume to evaluate early structural neurodegenerative changes that occurs in APOE ε4 positive elderly individuals who are at increased risk of developing LOAD. Furthermore, the evidence supports serial measurements and comparison of hippocampal volume based on age group, to track the progression of neurodegeneration in APOE ε4 carriers. Additional longitudinal studies are necessary to confirm whether the combination of MRI-detected hippocampal atrophy with APOE ε4 carrier status, can better predict the development of LOAD in cognitively normal individuals.

## 1. Introduction

Alzheimer’s disease (AD) is the commonest type of dementia and it is postulated to cause 60– 80% of cases worldwide (WHO, 2020; Alzheimer’s Association [AA], 2020). While older age group is recognized as the greatest risk factor for AD (AA, 2020; Rivan et al., 2020; Ibrahim et al., 2019), there are multiple other factors that are responsible for cognitive decline (Hussin et al., 2019; Lee et al., 2012; Shahar et al., 2013; Meramat et al., 2015). One of the most important non-modifiable risk factors is the apolipoprotein E (APOE) genotype which has been implicated in late-onset Alzheimer’s disease (LOAD). APOE is a 34 kDa glycoprotein containing 299 amino acid residues (Utermann, 1975) which functions as an essential constituent of plasma lipoproteins. APOE ε2 is a less common isoform and is considered protective against the development of AD (Wu and Zhao, 2016). Conversely, some studies postulate that ε2 may also be an important risk factor for AD (Kaur et al., 2005; van Duijn et al., 1995). The APOE ε3 allele is the most common genotype in the general population and apparently does not affect the risk of AD (Qiu et al., 2019). Universally, the APOE ε4 allele is the strongest identified genetic risk factor for both LOAD (Corder et al., 1993; Strittmatter et al., 1993) and sporadic AD (Poirier et al., 1993). APOE ε4 is usually present in 13.7% of worldwide population, but its frequency in LOAD patients is drastically increased to 40% (Farrer et al. 1997).

The pathological hallmarks underlying AD include Aβ plaques and neurofibrillary tangles (NFTs), and abnormal tau protein concentrations in the central nervous system (Sperling et al., 2011). Biomarkers indicating brain Aβ depositions are typically demonstrated as decreased levels of cerebrospinal fluid (CSF) Aβ42 and increased retention of amyloid tracer on positron emission tomography (PET) (Sperling et al., 2011; Suppiah et al., 2019). Structural magnetic resonance imaging (MRI) changes are also used as neurodegeneration biomarkers of AD, and known to manifest later compared to changes demonstrated on PET (Sperling et al., 2011). This suggests that the role of measuring structural changes may be limited in its ability to detect early changes in AD, prior to symptoms becoming apparent. A novel approach is to investigate the presence of APOE ε4 and combine the outcome with brain MRI detected changes in those who are at a higher risk of developing AD by the virtue of their age, family history, or other related risk factors. This can lead to improved diagnostic accuracy and better prediction of future clinical outcome with regards to LOAD. This stems from the compelling body of evidence that point to the crucial role of APOE ε4 in modulating cerebral Aβ and tau protein levels (Castellano et al., 2011; Strittmatter et al., 1994), neuronal activity (Sheline et al., 2010; Zhu et al., 2018), and ultimately gray matter volume (GMv) (Cacciaglia et al., 2018). Indeed previous systematic reviews and meta-analyses have explored the association between the APOE ε4 allele and brain structural changes; however these studies have only focused on selected regions of the brain, particularly the temporal lobe structures, with essentially no details on the subcortical regions (Liu et al., 2015; Cherbuin et al., 2007). Furthermore, these studies have placed a cap on the age group of the subjects when selecting their sample population as a criterion for inclusion or exclusion of subjects, effectively excluding younger age from adult groups. Therefore, this systematic review summarizes current available evidence on (1) the significant cortical and subcortical MRI-detected brain changes in neurodegeneration, (2) the relationship between the presence of APOE ε4 allele and the changes that occur in the cortical and subcortical brain structures, and (3) the association between APOE ε4 and cortical and subcortical changes examined with brain MRI across all age groups.

## 2. Material and methods

### 2.1 Sources of information and search approach

The following electronic databases were initially searched till August 19, 2019: Pubmed (http://www.ncbi.nlm.nih.gov/pubmed/), Scopus (https://www.scopus.com/home.uri), Ovid (http://www.ovid.com/site/index.jsp), and Cochrane (https://www.cochranelibrary.com/search). The search was updated on 3^rd^ March, 2020. Further, a review of included studies in the systematic review undertaken by Cherbuin et al. (2007) was conducted. The search was further complemented with a search of the grey literature. Additionally, cross referencing of all the bibliography of the primary sourced eligible articles was undertaken to identify the articles that fulfilled the inclusion criteria. The search method was in accordance with the Patient, measurement Instrument, Comparison, Outcome (PICO)-framework. The following search terms were entered: (“Apolipoprotein” OR “APOE”) AND (“brain imaging” OR “magnetic resonance imaging” OR “MRI”) AND (“Temporal lobe” OR “Hippocampus” OR “Medial Temporal Lobe” OR “Parahippocampal gyrus” OR “Amygdala” OR “Entorhinal cortex” OR “Parietal lobe” OR “Frontal lobe” “Occipital lobe” OR “Precuneus” OR “Entorhinal cortex” OR “Atrophy”) AND (“Brain size” OR “Brain volume” OR “Gray matter volume” OR “Cortical Thickness”).

### 2.2 Study selection

The articles had to fulfil the following inclusion criteria adopted from Cherbuin et al. (2007): (1) human subjects; (2) healthy subjects and/ or subjects with AD or dementia (including vascular dementia [VaD], dementia with Lewy bodies [DLB] and frontotemporal dementia [FTD]); (4) all subjects were examined using structural brain MRI technique; (5) brain regions were investigated for whole or regional alterations, and measurements made pertaining to GM thickness and/or GMv; (6) articles published in English language, and (7) full-text available articles. Articles that did not meet each of the listed criteria were excluded.

### 2.3 Eligibility Criteria

Assessment for eligibility was undertaken by conducting a screening of all articles obtained in accordance with the listed inclusion and exclusion criteria. Assessment for eligibility was undertaken by A.D.P. A.D.P is a PhD candidate working on ‘neuroimagenomics’ of MCI/AD patients. After identifying potentially eligible articles, duplicates were removed, and the first phase of the screening was performed based on title and abstract. When an abstract is deemed to provide inadequate information, a retrieval of the whole text was undertaken. The next phase involved the retrieval of full text articles which were evaluated to ensure that they meet the inclusion criteria. Consensus was often reached when there is concern regarding the inclusion or exclusion of an article.

### 2.4 Data information

The following information were extracted from each included article and presented in Tables 1 and 2. In Table 1, the following items are listed: (1) authors, (2) magnetic fields, (3) MRI parameters, (4) pulse sequences, (5) measurement and analysis method. Table 2 shows (1) authors, (2) age and gender of patient group, (3) age and gender of the control group, (4) main findings, and (5) remarks.

### 2.5 Risk of bias in individual studies

In order to determine the methodological quality of each eligible study, the Newcastle–Ottawa Scale (NOS, http://www.ohri.ca/programs/clinical_epidemiology/oxford.asp) designed purposely for case–control and cohort studies was used (Table 3). The NOS is apportioned into three categories: selection, comparability, and exposure/outcome. The maximum score on the NOS is nine points, which represents the highest methodological quality. To avoid bias, two reviewers (A.D.P. and M.M) independently conducted the methodological quality assessment. After quality assessment, comparison was made and consensus was reached when differences occurred in the scores. One point is awarded for each item under the Selection and Exposure/Outcome categories, while two points are awarded for the Comparability category. Regarding the response rate, a subcategory under Exposure, this was replaced because it did not fit the assessment criteria for the study. Therefore, this subcategory was replaced with item 9 to assess for ‘MRI data quality’ (Coppieters et al., 2016). This was to check whether researchers did perform visual inspection of ‘MRI data quality’. Overall, the maximum achievable score of a study was 9 based after modifying the NOS, which represented the test methodological quality. The NOS Scale is described in Table 3 as a footnote.

Regarding the study design and methodological quality, a level of evidence (LOE) was assigned to each study, in accordance with the 2005 classification system of the Dutch Institute for Healthcare Improvement (CBO) (http://www.cbo.nl/Downloads/632/bijlage_A.pdf) (Table 4). Following the clustering of studies with comparable methods or results, the strength of conclusion was determined (Table 5) based on the CBO classification, which accounted for the study designs as well as the risk of bias.

## 3.0 Results

### 3.1 Study Selection

The study selection process of studies that met the inclusion criteria is shown in Figure 1. Briefly, the initial search yielded a total of 800 studies. After eliminating duplicates, 475 studies remained. Further exclusion of studies based on titles and abstracts led to 239 articles remaining. After assessing the full-text articles, a total of 124 articles were removed, leaving total of 26 studies 115 which were included in the current study (Figure 1).

### 3.2 Study Characteristics

Briefly, from Table 1, for the studies that documented the MRI parameters, the magnetic field strengths ranged between 0.22 T to 7.0 T, repetition time (TR) and echo time (TE) varied between 2.73 to 10,000 ms and 2.0 to 30,153 ms respectively. Other parameters such as FA and ST varied between 7^0^ to 15^0^ and 1 to 10 mm respectively. While most studies acquired 3D T_1_ -weighted imaging using gradient echo pulse sequence, a few studies acquired T_2_-weighted imaging (Barboriak et al., 2000; Doody et al., 2000; Kerchner et al., 2014; Mueller and Weiner (2009); Mueller et al., 2008; Donix et al., 2013). Even though most studies used volumetric measures for analysis on semi-or fully automated software, a small number of studies used manual tracing (Du et al., 2000; Tanaka et al., 1998; Geroldi et al., 2000; Geroldi et al., 1999; Lu et al., 2011; Juottonen et al., 1998; Barzokis et al., 2006), and visual analysis (Cotta Ramusino et al., 2019; Barber et al., 1999; Doody et al., 2000).

### 3.3 Risk of bias and level of evidence

Table 3 shows the results of the methodological quality assessment. The LOE is graded as follows: -, score not fulfilled; +, score fulfilled; /, answer is unclear. Methodological quality varied from poor to good, between 3 of 9 and 8 of 9. Most studies lost points on ‘selection of controls’, and ‘definition of controls’, and ‘MRI data quality’. This was mainly because authors did not provide the required information or the information was inadequate. Most eligible studies matched participants for age and gender. A few studies controlled for additional factors (e.g. APOE ε4).

In addition, Table 2 shows the characteristics of each study. All studies in the review were assigned an LOE B, due to the inclusion of only case–control and cohort type of studies.

### 3.4 Syntheses of Results

Overall, almost 87% of the eligible articles investigated the temporal lobe, yielding a total of 100 articles. A total of 22 articles investigated the frontal lobe, while the parietal lobe was investigated in 18 articles. The occipital lobe was the least studied, and was found to be investigated in only 7 articles. Several other cortical and subcortical regions were investigated in a total of 29 studies.

#### 3.4.1 Frontal lobe

From Table 2, most studies reported that APOE ε4 carriers demonstrated reduced GMv and/ or increased rate of atrophy of the frontal lobe as a whole or its specific regions which included the following: left frontal lobe. However, in an APOE ε4 dose-dependent manner, the evidence regarding the direction of change of frontal lobe GMv are conflicting. A few studies also found APOE ε4 carriers to demonstrate thinner cortex in the following regions: bilateral lateral frontal, left rostral midfrontal, right caudal midfrontal regions, superior frontal gyrus, and dorsolateral frontal regions. This contrasts with studies that found increased cortical thickening of the lateral medial orbitofrontal cortical region (Chang et al., 2016), and dorsolateral frontal region (Sampedro et al., 2015; Fan et al 2010). The left side region of the dorsolateral region was found to show increased cortical thickening compared to the right, as indicated by Fan et al. (2010). Several studies did not identify an effect of APOE ε4 on either the frontal lobe size, or on the side of the caudal and rostral middle, superior, inferior, and orbitofrontal regions. There is evidence of the lack of an association between APOE ε4 and frontal lobe atrophy, and several of its regions i.e. right superior frontal gyrus, the lateral and medial frontal region. Nevertheless, an association was found between APOE ε4 and reduced GMv in the inferior frontal lobe (Chen et al 2012), and the dorsolateral frontal region (Sampedro et al., (2015). Another study postulated that there was no association between APOE ε4 and cortical thinning of the inferior frontal cortex (Sabuncu et al., 2012).

Moderate evidence shows reduced middle frontal gyrus size among APOE ε4 carriers (strength of conclusion 2). Some evidence showed cortical thinning in the lateral frontal, left rostral midfrontal, right caudal midfrontal, and superior frontal gyrus in APOE ε4 carriers (strength of conclusion 3). Additionally, there is some evidence of increased cortical thickening of the lateral medial orbitofrontal cortical region (strength of conclusion 3). Moderate evidence showed that there is no APOE ε4 effect on the frontal lobe (strength of conclusion 2), while there is some evidence on the lack of influence of APOE ε4 on specific regions of the frontal lobe, specifically the caudal and rostral middle, superior, inferior, and orbitofrontal regions (strength of conclusion 3). There is moderate evidence of the lack of association between APOE ε4 and the frontal lobe (strength of conclusion 2).

#### 3.4.2 Parietal lobe

From the studies, APOE ε4 carriers were found to demonstrate cortical thinning in the medial, lateral parietal, inferior parietal, precuneus, left parietal gyrus, superior parietal gyrus, angular gyrus, and the right hemisphere of the parietal lobe. Contrarily, APOE ε4 carriers were reported in other studies to demonstrated increased cortical thickness in the precuneus, inferior parietal region, and the right parietal regions. Although there is contradictory evidence regarding the direction of volumetric change in the parietal lobe in APOE ε4 carriers, its regions such as the right angular gyrus and precuneus were found to be reduced. Furthermore, there is conflicting evidence regarding the association between APOE ε4 and parietal atrophy; however some reports show association between APOE ε4 and the inferior and superior parietal cortices. There is abundant evidence pointing to an association between APOE ε4 status and increased rates of cortical thinning in the precuneus among MCI patients. Contrarily, there was a report of an absence of a significant association between APOE ε4 and the size of the inferior parietal cortex and sulcus (Sabuncu et al., 2012). There are conflicting evidence regarding the effect of APOE ε4 on the parietal lobe cortical region.

In conclusion, there is evidence that APOE ε4 carriers show cortical thinning in the medial lateral parietal and left parietal gyrus (strength of conclusion 3). There is moderate evidence that APOE ε4 carriers demonstrate cortical thinning in the superior parietal gyrus (strength of conclusion 2). There is moderate evidence that the cortical thickness of the precuneus changes in APOE ε4 carriers (strength of conclusion 2). However, inconclusive evidence exists regarding the direction of change of cortical thickness in the precuneus, and inferior parietal region in APOE ε4 carriers (strength of conclusion 4). There is moderate evidence of GM changes in the parietal lobe in APOE ε4 carriers (strength of conclusion 2). There is moderate evidence that GMv in the precuneus is reduced in APOE ε4 carriers (strength of conclusion 2).

#### 3.4.3 Temporal lobe

##### 3.4.3.1 Hippocampus

Several studies consistently reported that APOE ε4 carriers exhibited decreased hippocampal GMv or cortical thinning and/ or observed association between APOE ε4 and reduced hippocampal GMv. An interesting point is that apart from these studies engaging older adults the majority of these studies also involved MCI and/ or AD patients with or without healthy controls (HCs). Several studies did not find any effect of APOE ε4 on reduced hippocampal GMv or size reduction rate. However, there is evidence of increased APOE ε4 dose effects on hippocampal atrophy. Several studies did not find an association between APOE ε4 dose effect and reduced hippocampal GMv or cortical thinning measurements. However a study (Kerchner et al., 2014) found an association between APOE ε4 dose effect on cortical thinning of the hippocampal subregions.

In conclusion, moderate evidence shows that the hippocampal volume is not only reduced in APOE ε4 carriers but it is also associated with APOE ε4 (strength of conclusion 2). Furthermore, there is moderate evidence of reduced cortical thickness or GMv of some hippocampal subfields (CA1-3, dentate gyrus, and subiculum) (strength of conclusion 2). Some evidence shows that APOE ε4 dose effect is associated with cortical thinning of the hippocampal subregions (strength of conclusion 3).Moderate evidence shows that APOE ε4 has no effect on hippocampal atrophy or rate of atrophy (strength of conclusion 2). However there is moderate evidence that increased APOE ε4 dose effects is observed on hippocampal atrophy (strength of conclusion 2). Further, moderate evidence shows the lack of association between APOE ε4 dose effect and hippocampal atrophy or volume (strength of conclusion 2). There is moderate evidence of decreased cortical thickness in the hippocampal subregions (strength of conclusion 2). There is evidence to show that APOE ε4 exerts no significant effect on hippocampal thickness (strength of conclusion 3).

##### 3.4.3.2 Amygdala

Several studies reported consistent reduction of volumetric measures, or atrophy, and increased atrophy rate of the amygdala in APOE ε4 carriers. These studies suggest age-related atrophy of the amygdala similar to what was observed in the hippocampus. There are conflicting reports regarding the effect of APOE ε4 on amygdala volume. Furthermore, reports on amygdala asymmetry in APOE ε4 carriers are conflicting.

In conclusion, there is moderate evidence of reduced amygdala volumes in older individuals with MCI and AD compared to HCs (strength of conclusion 2). However, there is inconclusive evidence regarding the effect of APOE ε4 on amygdala volumes or its atrophy, and symmetrical differences in demented and non-demented older adults (strength of conclusion 4).

##### 3.4.3.3 Entorhinal Cortex

Consistently, APOE ε4 carriers were found to exhibit cortical thinning (or reduced volume) in the entorhinal cortex. However, one study found the opposite which was observed in younger participants (DiBattista et al., 2014). Furthermore, reports of effect or an association between APOE ε4 and entorhinal cortical atrophy/thinning or its rate of reduction are conflicting. Interestingly, a single study observed an association between APOE ε4 and increased entorhinal cortical thickening among those with early MCI and HCs (Li et al., 2017).

In conclusion, moderate evidence shows that APOE ε4 carriers demonstrate cortical thinning in the entorhinal cortex (strength of conclusion 2). There is indistinct evidence regarding the effect of APOE ε4 and likewise the association between APOE ε4 and entorhinal size reduction (strength of conclusion 4).

##### 3.4.3.4 Parahippocampal gyrus

Parahippocampal gyrus was found to demonstrate decreased cortical thickening or increased atrophy or rate of reduction among APOE ε4 carriers who were within the older age bracket. No evidence of APOE ε4 effect was observed on the parahippocampal gyrus. There is strong evidence to support that APOE ε4 carrier status is associated with reduced GMv (or thickness) or increased rate of atrophy in the parahippocampal gyrus. Interestingly, one study found an association between APOE ε4 and increased cortical thickening in the parahippocampal gyrus in healthy participants and those with early MCI.

In conclusion, moderate evidence shows that APOE ε4 carriers demonstrate reduced cortical thickening, increased atrophy, or increased rate of atrophy in the parahippocampal gyrus (strength of conclusion 2). Further, moderate evidence shows no APOE ε4 effect on parahippocampal gyrus (strength of conclusion 2). Additionally, moderate evidence shows that APOE ε4 is associated with reduced volume and increased rate of atrophy of the parahippocampal gyrus (strength of conclusion 2).

##### 3.4.3.5 Other temporal lobe regions of interest

An interesting point to highlight is the report of increased cortical thickness in APOE ε2 carriers relative to APOE ε4 carriers (Konishi et al., 2016; Groot et al., 2016). This suggests that APOE ε2 may offer some protective role against dementia in both young adults older age group.

In conclusion, from Table 2, there is reasonable evidence that APOE ε4 carriers demonstrate reduced GMv in the middle temporal lobe, temporal pole, and in general, the whole temporal lobe (strength of conclusion 2). Further, some evidence shows that APOE ε4 carriers demonstrate reduced cortical thinning in the middle temporal gyrus, and medial and temporal regions; and GMv reduction or rate of reduction in the fusiform, temporoparietal and occipitotemporal regions (strength of conclusion 3). Conversely, some evidence shows that male APOE ε4 carriers demonstrate increased cortical thickening in the temporoparietal regions (strength of conclusion 3). Furthermore, there is moderate evidence that APOE ε4 is associated with increased rate of cortical thinning in temporoparietal cortex, and that APOE ε4 does not exert an effect on temporal lobe volume (strength of conclusion 2). There is also some evidence that APOE ε4 is associated with lower cortical thickening of the temporal lobe cortex and reduced GMv in the fusiform gyrus and temporal pole cortex (strength of conclusion 3).

#### 3.4.4 Occipital lobe

APOE ε4 carriers were reported to demonstrate increased GMv in the left middle occipital cortex in a dose-dependent manner, with evidence of increased rate of atrophy in bilateral lingual gyri in APOE ε4 carriers. Increased cortical thickening was found in the occipital region, and in the right hemisphere in the occipital region specifically in male APOE ε4 carriers. On the other hand, the occipital region was reported to demonstrate cortical thinning in APOE ε4 carriers. One study found no association between APOE ε4 and atrophy of the occipital lobe (Doody et al., 2000). A single study found an association between APOE ε4 and decreased volume of right lingual gyrus (Tosun et al., 2010), and another found an association between APOE ε4 and lower volumes of the left cuneus (Yokoyama et al., 2015).

In conclusion, although there is evidence of increased GMv or cortical thickness in some cortical regions of the occipital lobe in APOE ε4 carriers, only the bilateral lingual gyri were found to demonstrate atrophy APOE ε4 carriers (strength of conclusion 3). This is reflected in the differences in the association between APOE ε4 and occipital lobe or its regions.

#### 3.4.5 Other cortical gray matter regions

In conclusion, based on the evidence presented in Table 2, it is clear that there is some evidence that APOE ε4 carriers showed decreased GMv in the insula, and several regions of the cingulate cortex (strength of conclusion 3). Further, some evidence shows an association between APOE ε4 and reduced GMv in the anterior cingulate, cerebral cortex; and between APOE ε4 and increased cortical thickness in the right caudal anterior cingulate cortex (strength of conclusion 3). Also, there is some evidence that APOE ε4 has no effect on the cortical thickness of bilateral cingulate cortices in AD patients (strength of conclusion 3). However, there is some evidence of additive effect of APOE ε4 on reduced GMv in the right cerebellar crus (strength of conclusion 3). There is moderate evidence of reduced GMv and lower cortical thickening in the posterior cingulate cortex (PCC) in APOE ε4 carriers (strength of conclusion 2). There is some evidence that APOE ε4 exerts an effect on cerebral cortex atrophy (strength of conclusion 3). However there is lack of association between APOE ε4 and cerebral volume (strength of conclusion 2), and cerebellar GMv reduction (strength of conclusion 3).

#### 3.4.6 Other subcortical gray matter regions

In summary, based on the findings shown in Table 2, APOE ε4 may have minimal to no effect on the size of the thalamus from young to middle age, while in older adults, it may exert more negative effect (strength of conclusion 3). Further, some evidence shows that older adult APOE ε4 carriers with MCI exhibit increased GM atrophy of the thalamus (strength of conclusion 3). There is some evidence of increased rate of subcortical atrophy of the putamen and loss of GM in the right caudate nucleus in APOE ⍰4 carriers (strength of conclusion 3). Inconclusive evidence exists regarding the association between APOE ⍰4 carriers and basal ganglia GMv (strength of conclusion 4). Some evidence shows subcortical atrophy in the accumbens, right ventral striatum and sulcal widening among APOE ε4 carriers (strength of conclusion 3).

## 4.0 Discussion

Several neuroplastic changes were found to occur not only in cortical areas of GM, but several subcortical regions in individuals carrying the APOE ε4 allele relative to noncarriers. These changes have often manifested as reduced GMvs or cortical thinning in both cortical and subcortical structures. Patterns of GM thinning is thought to be reflective of synaptic pruning size alterations and number of glia or neuronal size (Huttenlocher, 1979; Cotter et al., 2002). GMv reduction in the middle frontal gyrus may provide better insight regarding the underlying pathophysiological changes that occur in APOE ε4 carriers, compared to changes in the right lateral orbitofrontal cortex or lateral prefrontal cortex. MRI studies reveal that to some extent, the cortical thinning in the frontal lobe subcortical structures i.e. the lateral frontal, left rostral midfrontal, right caudal midfrontal, and superior frontal gyrus; and increased cortical thickening in the lateral medial orbitofrontal cortical region in APOE ε4 carriers may be of clinical significance. Overall, it appears that the cortical thickness measurements of the frontal lobe subcortical structures may be able to elicit more neuroplastic changes compared to GMv measurements.

Decreased volumes in specific parietal lobe subcortical structures i.e. right angular gyrus, may provide valuable details about structural changes in APOE ε4 carriers. We noted that the cortical thinning in the superior parietal gyrus appear to better reflect the effect of APOE carrier status with neurodegeneration. The NIA-AA guidelines recommend that the presence of cortical thinning or GMv loss in the lateral and medial parietal gyri is a marker of AD-pathophysiology (Sperling et al., 2011). Nevertheless, we did find strong evidence to support medial and lateral parietal lobe atrophy in the APOE carriers. In fact, Mattson et al (2018) detected increased cortical thickness of these regions among the APOE positive carriers as compared to non-carriers. Similar findings have been noted by Chételat et al. (2010) and Johnson et al.,2014, whereby amyloid positive individuals demonstrated increased GMv in the temporal and lateral parietal regions, which was postulated to be due to brain swelling associated with glial activation in preclinical AD stages (Chételat et al., 2010; Johnson,2014). There is consistent reports of precuneal GMv reduction and its association with APOE ε4. The precuneus together with the PCC has been the main focus of several studies investigating cognitively normal individuals and/or AD because it is recognized as one of the key regions that demonstrates cortical thinning (Lehmann et al., 2010), hypometabolism on ^18^F-fluorodeoxyglucose (FDG)–PET (Minoshima et al., 1997), lower levels of N-acetyl aspartate/creatine, elevated myoinositol/Cr (Voevodskaya et al., 2016; Suri et al., 2017), and histopathologic changes (Braak and Braak, 1991) in the early course of AD. This may suggest that precuneal GM volumetry may better provide insight on neuronal injury compared to cortical thickness measures.

There is strong evidence showing that APOE ε4 carriers demonstrate reduced cortical thickening or increased GMv reduction in medial temporal lobe regions (hippocampus, amygdala, entorhinal cortex, and parahippocampal gyrus) widely known to play significant roles in memory and implicated in neural correlates of AD. An important highlight to the reduction of cortical thickening in the hippocampus is the involvement of its subfields (CA1-3, dentate gyrus, and subiculum) which appear to be largely driven by old age. There are proponents of the hippocampal subfield volumetric measures in place of the traditional whole hippocampal volumetry as AD biomarker (Maruszak and Thuret, 2014; La Joie et al., 2013). This stems from evidence of differences in vulnerability of the hippocampal subfields to AD neuropathology (Braak and Braak, 1991; Braak and Braak, 1993). The findings on the effect and association between APOE ε4 and amygdala, entorhinal cortex, and parahippocampal gyrus is mixed, suggesting their limitation as potential AD biomarkers.

The occipital lobe is the least studied, possibly due to the reason that visual symptoms are uncommon and that this region is not usually affected until relatively late in the neocortical stages of AD (Smith et al. 2001). From the findings, it seems plausible to suggest that the whole occipital lobe may better provide insight regarding structural changes among APOE ε4 carriers compared to its subcortical regions. The PCC may be an important structure to earmark for analysis during the pathophysiological process of AD or in healthy individuals with APOE ε4.

To put these results into a broad perspective, these findings expand our knowledge that apart from cortical structures widely touted as being influenced by APOE ε4, subcortical structures especially that of the medial temporal lobe are also vulnerable and influenced to a large extent by advanced age, and APOE ε4 carriership.

### 4.1 Strengths and Limitations

Compared with previous systematic reviews and meta-analysis (Liu et al., 2015; Cherbuin et al., 2007), the current study did not restrict the inclusion criteria to the number of sample sizes, age of participants, and regional structures investigated in each study. Further, a level of evidence B was assigned because comparative studies including patient-control studies and cohort studies were included. However, the studies identified showed great variation in a number of variables, i.e. the magnetic field strength, MRI parameters, measurement and analytic methods, region(s) of interest studied, disproportionate distribution of sample size and gender, age range or average age, and differences in measuring tools applied for determining cognitive status of participants. It is possible that these variations to some extent may have led to a few of the discordant findings reported. Nonetheless, the results are striking in that there is much consistency in structural alterations in APOE ε4 carriers.

## 5.0 Conclusions

The present data supports analyzing the hippocampal GM volume in particular as part of the diagnostic process in healthy individuals with APOE ε4 and those at risk of developing AD. Further the study supports the use of hippocampal atrophy as an invaluable AD biomarker that can contribute to tracking progressive structural brain alterations different age group who are APOE ε4 carriers. However, further longitudinal studies may be necessary to confirm whether a combination of both hippocampal atrophy and APOE ε4 will be capable of predicting the development of AD in healthy individuals.

## Data Availability

Data available within the manuscript.

## Author contributions

Conception and study design (ADP, MM), data collection (ADP), interpretation of results (ADP, MM, and SS); drafting the manuscript work or revising it critically for important intellectual content (ADP, MM, SS, and NFR); approval of final version to be published and agreement to be accountable for the integrity and accuracy of all aspects of the work (ADP, MM, SS, and NFR).

## Compliance with Ethical Standards

### Funding

None

### Ethical approval

All procedures performed in studies involving human participants were in accordance with the ethical standards of the institutional and/or national research committee and with the 1964 Helsinki declaration and its later amendments or comparable ethical standards.

### Informed consent

Informed consent was obtained from all individual participants included in the studies reviewed in this paper.

---

Figure 1. PRISMA flowchart of the study selection process

Table 1: MRI Parameters and Measurement Techniques

Table 2. Evidence table of studies included in qualitative synthesis Table 3 Methodological quality of included studies

Table 3 Methodological quality of included studies

Table 4 LOE, according to the 2005 classification system of the Dutch Institute for Healthcare Improvement CBO

Table 5 Strength of Conclusion (modified table)

