## Supplemental Tables for "Apolipoprotein E genotype and MRI-detected brain alterations pertaining to neurodegeneration: A systematic review"

Table 1: MRI Parameters and Measurement Techniques

| Authors | Magnet strength (T) | TR/TE/TI (ms) | FA | ST | Pulse Sequence | Measurement and Analysis Method |
| --- | --- | --- | --- | --- | --- | --- |
| Cacciaglia et al. (2018) | 3.0 | 6.16/2.33/450 | 12^0^ | - | Fast SPGR | Volumetric (SPM) |
| Groot et al. (2018) | 3.0 or 1.5 | - | - | - | 3D T_1_-weighted; T_2_*-weighted | Volumetric (SPM) |
| Kirsebom et al. (2018) | 3.0 | 4.5/2.2/853 | 8^0^ | 1.2 | 3D T_1_-weighted TFE; MP-RAGE | Volumetric (FreeSurfer) |
| Koval et al. (2018) | - | - | - | - | 3D T_1_-weighted | Cortical Thickening |
| Wang et al. (2018) | 1.5 | 15/7/-  6000/100/1900 | 15^0^ | - | 3D T­_1_-weighted FFE  FLAIR | Manual Tracing and Semiautomatic |
| Wang et al. (2019) | 1.5 | - | - | - | - | Volumetric |
| Mattsson et al. (2018) | 3.0 | 1950/3.4/- | - | - | 3D MP-RAGE | Volumetric and Cortical Thickening (FreeSurfer) |
| Moon et al. (2018) | 1.5 | 15/5/- | 20^0^ | 1.5 | 3D T_1_-weighted | Volumetric and Shape |
| Rane et al. (2018) | 3.0 | 2300/2.98/900 | - | 1-1.2 | - | Volumetric |
| Sundermann et al. (2018) | 1.5 | - | - | - | - | Volumetric (FreeSurfer) |
| Dong et al. (2019) | 3.0 | 33/5 | 30^0^ | 1.5 | 3D T_1_-weighted SPGR | Surface (FSL) |
| Barboriak et al. (2000) | 1.5 | 4000/15, 105 | - | 2.5 | T_2_-weighted FSE | Visual Analysis |
| Lehtovirta et al. 1995 | 1.5 | 10/4/250 | 12^0^ | 1.5-1.8 | 3D MP-RAGE | Volumetric Analysis/Manual Tracing |
| Du et al. (2000) | 1.5 | 10/7/300 | 15^0^ | 1.4 | 3D MP-RAGE | Manual Tracing |
| Barber et al. (2000) | 1.0 | 11.4/4.4/400 | - | 1.0 | 3D MP-RAGE | Volumetric Analysis/Manual Tracing |
| Boccardi et al. (2004) | 1.5 | 10/4/300 | 10^0^ | 1.3 | 3D T_1_-weighted GRE | Volumetric Analysis |
| Reiman et al. (1998) | 1.5 | 33/5 | 30^0^ | 1.5 | 3D T_1_- weighted SPGR | Volumetric Analysis/Manual Tracing |
| Schmidt et al. (2008) | 1.5 | 600/30 | - | 5.0 | T_1_-weighted | Volumetric Analysis/Manual Tracing |
| Basso et al. (2006) | 1.5 | 24/5 | 45^0^ | - | 3D SPGR | Volumetric Analysis/Manual Tracing |
| Espeseth et al. (2006) | 1.5 | 2730/3.43/1000 | 7^0^ | - | 3D MP-RAGE | Volumetric and Cortical Thickness Analysis |
| Tanaka et al. (1998) | 0.22 | 500/30 | - | 10.0 | T­_1_-weighted | Manual Tracing |
| Bigler et al. (2002) | 0.5 | 500/15 | - | 5 | T­_1_-weighted | Volumetric Analysis |
| Barber et al. (1999) | 1.0 | 11.4/4.4/400 |  | 1 | 3D T_1_-weighted | Visual Rating |
| Carmelli et al. (2000) | 1.5 | 2000/20 or 100 | - | 5 | CSE Double-Echo | Semiautomated Segmentation Analysis |
| Pennanen et al. (2006) | 1.5 | 9.7/4 | 10^0^ | 2 | 3D T_1_-weighted GRE | Voxel-based Morphometry (Statistical Parametric Mapping) |
| Morra et al. (2009) | 1.5 | 2400/-/1000 | 8^0^ | - | 3D MP-RAGE | Volumetric and Surface-based Analysis |
| Goltermann et al. (2019) | 3.0 |  |  |  | - | Volumetric and Surface Analysis |
| Konishi et al. (2018) | 3.0 | 2300/2.08 | 9^0^ | - | 3D MP-RAGE | Voxel-based Morphometry |
| Kelly et al. (2018) | 3.0  3.0 | 3.9/9.5  9.6/3.9 | 12^0^  12^0^ | 1  1 | SPGR | Volumetric Analysis (FreeSurfer) |
| Alemany et al. (2018) | 1.5 | 11.9/4.2 | 15^0^ | 1.2 | Fast SPGIR | Volumetric Analysis (FreeSurfer) |
| Tosun et al. (2010) | 1.5 | 9/4 | 8^0^ | - | 3D MP-RAGE | Volumetric and Thickness Analysis |
| Hua et al. (2008) | 1.5 | 2400/-/1000 | 8^0^ | - | 3D MP-RAGE | Volumetric Analysis |
| Filippini et al. (2009) | 1.0 | 20/5  11.4/4 | 30^0^  8^0^ | 1.3  1.3 | 3D T_1_-weighted | Volumetric Analysis |
| Jak et al. (2007) | 1.5  1.5 | - | - | 1.2 | SPGR | Volumetric Analysis |
| Doody et al. (2000) | 1.5 | 3500/90 | - | 5 | T_2_-weighted | Visual Analysis |
| Geroldi et al. (2000) | 1.5 | - | - | - | 3D MP-RAGE | Manual Tracing |
| Mishra et al. (2018) | 1.5  3.0 | - | - | - | 3D MP-RAGE | Volumetric and Thickness Analysis (FreeSurfer) |
| Bussy et al. (2019) | 3.0 | - | - | - | - | Volumetric Analysis (FreeSurfer) |
| Taylor et al. (2014) | 1.5 | 9/2 | 15^0^ | - | 3D Fast SPGR | Volumetric Analysis |
| Kerchner et al. (2014) | 7.0 | 5 to 6s/49 | - | - | T_2_-weighted FSE | Cortical Thickening Analysis |
| Fennema-Notestine et al. (2011) | 1.5 | 2730/3.31/1000 | 7^0^ | 1.33 | 3D MP-RAGE | Volumetric and Thickness Analysis |
| Andrawis et al. (2012) | 1.5 | - | - | - | - | Volumetric Analysis |
| Chang et al. (2016) | 3.0 | - | - | - | 3D MP-RAGE | Volumetric and Thickness Analysis |
| Honea et al. (2009) | 3.0 | 2500/4.38/1100 | 8^0^ | - | 3D MP-RAGE | Volumetric Analysis (Statistical Parametric Mapping) |
| Chang et al. (2014) | 1.5 | - | - | - | - | Volumetric and Thickness Analysis (FreeSurfer) |
| Li et al. (2016) | - | - | - | - | - | Volumetric and Shape Analysis |
| Okonkwo et al. (2010) | 1.5 | - | - | - | 3D MP-RAGE | Volumetric Analysis (FreeSurfer) |
| Tosun et al. (2011) | 1.5 | 9/4 | 8^0^ | - | 3D MP-RAGE | Cortical Thickening Analysis |
| Tang et al. (2015) | 1.5 | - | - | - | - | Volumetric and Shape Analysis |
| Burggren et al. (2008) | 3.0 | -/3.7/500 | - | - | SPGR | Volumetric and Cortical Thickness Analysis |
| Reiter et al. (2017) | 3.0 | 9.5/3.9  9.6/3.9 | 12^0^  12^0^ | 1  1 | SPGR  SPGR | Volumetric Analysis |
| Lampert et al. (2014) | 1.5 | - | - | - | 3D MP-RAGE | Volumetric and Cortical Thickness Analysis (FreeSurfer) |
| Bender and Raz (2012) | 4.0 | 1600/4.38/800 | - | 1.34 | 3D MP-RAGE | Volumetric Analysis |
| Mueller and Weiner (2009) | 4.0 | 3500/0.019 | - | 2 | T_2_-weighted FSE | Volumetric Analysis |
| Mueller et al. (2008) | 4.0 | 2300/3/950  8390/70 | 7^0^  150^0^ | -  - | 3D MP-RAGE  T_2_-weighted | Volumetric Analysis (FreeSurfer) |
| Donix et al. (2010) | 3.0 | 2300/2.93 | - | 1 | 3D MP-RAGE | Volumetric and Cortical Thickening Analysis |
| Ferencz et al. (2013) | 1.5 | 15/7 | 15^0^ | 1.5 | 3D FFE | Volumetric Analysis |
| Spampinato et al. (2011) | 1.5 | - | - | - | 3D MP-RAGE | Voxel-based Morphometry (Statistical Parametric Mapping) |
| Ma et al. (2016) | 3.0 | 1900/3.44 | 9^0^ | 1 | 3D MP-RAGE | Volumetric Analysis |
| Lu et al (2011) | 1.5  1.5 | 24/7  25/5 | 35^0^  35^0^ | 1.5  1.4 | SPGR  SPGR | Manual Tracing |
| Dean et al. (2014) | 3.0 | - | - | - | IR-SPGR | Volumetric and Fraction Analysis |
| Wolk et al. (2010) | - | -- | - | - | - | Volumetric and Thickness Analysis |
| Soldan et al. (2015) | 1.5  1.5 | 24/2  24/3 | 20^0^  45^0^ | 2  1.5 | SPGR  SPGR | Volumetric and Thickness analysis |
| Fan et al. (2010) | 1.5 | 90/3.9 | 8^0^ | 1.2 | 3D MP-RAGE | Cortical Thickness Analysis |
| Banks et al. (2017) | - | - | - | - | - | Volumetric Analysis (FreeSurfer) |
| Walsh et al. (2013) | 1.5 | 27/9 | 25^0^ | 1.6 | SPGR | Volumetric and Manual Tracing Analysis |
| Li et al (2017) | - | - | - | - | 3D MP-RAGE/IR-SPGR | Thickness, Surface, and Volumetric Analysis (FreeSurfer) |
| Chen et al. (2012) | 1.5 | 33/5 | 30^0^ | 1.5 | SPGR | Voxel-based Morphometry (Statistical Parametric Mapping |
| Hostage et al. (2014) | - | - | - | - | - | Volumetric and Thickness Analysis (FreeSurfer) |
| Donix et al. (2013) | 3.0 | 5200/105 | - | 3.0 | T_2_-weighted FSE | Volumetric and Thickness Analysis |
| Shi et al. (2014) | - | - | - | - | - | Segmentation and Surface Reconstruction |
| Hafsteinsdottir et al. 2012 | - | - | - | - | FLAIR | Volumetric Analysis |
| Novellino et al. (2019) | 1.5 | 11.2/4.2/450 | 12^0^ | 1 | SPGR | Voxel-based Morphometry |
| Juottonen et al. (1998) | 1.5 | 10/4/250 | 12^0^ | 1.5-1.8 | 3D MP-RAGE | Manual tracing |
| Wilhelm et al. (2008) | 1.5 | - | - | 1.3 | FFE | Volumetric Analysis |
| Cherbuin et al. (2008) | 1.5 | 28.05/2.64 | 30^0^ | 2.0 | FFE | Manual measurement and  Voxel-based Morphometry (Statistical Parametric Mapping) |
| Biffi et al. (2010) | 1.5 | - | - | - | 3D MP-RAGE | Volumetric Analysis and Thickness (FreeSurfer) |
| Ystad et al. (2009) | 1.5  1.5 | 9.5/2.2/450  2.730/3.39/1000 | 7^0^  7^0^ | -  - | SPGR  3D MP-RAGE | Volumetric, Surface, and Thickness Analysis (FreeSurfer) |
| Schuff et al. (2009) | 1.5 | 9/4 | 8^0^ | - | 3D MP-RAGE | Volumetric Analysis (Semiautomated tracing) |
| Stewart et al. (2011) | 1.5 | - | - | - | - | Volumetric Analysis |
| Protas et al. (2013) | 1.5 | 33/5 | 30^0^ | 1.5 | SPGR | Volumetric Analysis (Semiautomated Algorithm) |
| Hoogendam et al. (2012) | 1.5 | 13.8/2.8/400 | 20^0^ | 1.6 | Fast SPGR | Volumetric Analysis (FreeSurfer) |
| Geroldi et al. (1999) | 1.5 | 10/4/300 | 10^0^ | - | 3D MP-RAGE | Manual tracing |
| Lehtovirta et al. (1996) | 1.5 | 10/4/250 | 12^0^ | 1.5-1.8 | 3D MP-RAGE | Volumetric Analysis (Standard Anatomical Atlas) |
| O’Dwyer et al. (2012) | 3.0 | 7.92/2.48 | - | 1 | 3D MDEFT | Volumetric Analysis (FSL) |
| Sabuncu et al. (2012) | - | - | - | - | - | Volumetric Analysis (FreeSurfer) |
| Bunce et al. (2012) | 1.5 | 8.93/3.57 | 8^0^ | 1.5 | FFE | Volumetric Analysis (FreeSurfer) |
| Hostage et al. (2013) | 1.5 | - | - | - | - | Volumetric Analysis (FreeSurfer) |
| DiBattista et al. (2014) | 3.0 | - | - | 1.25 | 3D MP-RAGE | Voxel-based Morphometry (DARTEL, Statistical Parametric Mapping) |
| Manning et al. (2014) | 1.5 | - | - | - | 3D MP-RAGE | Multi-Atlas Propagation and Segmentation |
| Khan et al. (2014) | 3.0 | - | - | - | 3D MP-RAGE | Volumetric Analysis (FreeSurfer) |
| Holland et al. (2013) | - | - | - | - | 3D T_1_ GRE | Volumetric Analysis (Quarc) |
| Lyall et al. (2013) | 1.5 | 10/4/500 | - | - | 3D MP-RAGE | Volumetric Analysis (FreeSurfer) |
| Morgen et al. (2015) | 1.5  1.5 | 9.3 to 20/3.93 to 4.38  9000-10000/100-110/2500 | 15^0^ | 1-1.2  5 to 6 | FLAIR  FLAIR | Volumetric Analysis (Statistical Parametric Mapping) |
| Risacher et al. (2015) | - | - | - | - | - | Volumetric Analysis (FreeSurfer) |
| Yokoyama et al. (2015) | 1.5 | - | - | - | FLAIR | Voxel-Based Morphometry (DARTEL) |
| Sampedro et al. (2015) | - | - | - | - | - | Cortical Thickness (FreeSurfer) |
| Khan et al. (2017) | 1.5/3.0 | - | - | - | 3D MP-RAGE | Volumetric Analysis (FreeSurfer) |
| Falahati et al. (2017) | 1.5 | - | - | - | 3D MP-RAGE | Volumetric Analysis (FreeSurfer) |
| Rogne et al. (2016) | 1.5 | 2300/4/1000 | 8^0^ | - | 3D MP-RAGE | Volumetric Analysis (NeuroQuant) |
| Konishi et al. (2016) | 1.5 | 22/10 | 30^0^ | - |  | Volumetric Analysis |
| Habes et al. (2016) | 1.5 | 1900/3.4 | 15^0^ | 1 | 3D MP-RAGE | Volumetric Analysis |
| Fang et al. (2019) | 1.5 | - | - | - | SPGR | - |
| Lupton et al. (2016) | - | - | - | - | - | Volumetric Analysis (FreeSurfer) |
| Hobel et al. (2019) |  |  |  |  |  | Volumetric Analysis (FreeSurfer and ITK-SNAP |
| Schreiber et al. (2017) | - | - | - | - | 3D MP-RAGE | Voxel-Based Morphometry (FreeSurfer) |
| Haller et al. (2017) | 3.0 | 2300/2.3 | - | - | 3D T_1_-weighted | Voxel-Based Morphometry (FreeSurfer) |
| Nao et al. (2017) | 3.0 | 9.5/4.6 | 20^0^ | 1.2 | 3D TFE | Voxel-Based Morphometry |
| Li et al. (2019) | 3.0 | - | - | - | - | Visual Reading |
| Hays et al. (2019) | - | 8/3.1/600 | 12^0^ | - | SPGR | Volumetric and Cortical Thickening Analysis |
| Herrmann et al. (2019) | 3.0 | 2300/2.27 | - | 1 | 3D MP-RAGE | Volumetric Analysis (FSL) |
| Foley et al. (2017) | - | 8/3/450 | 20^0^ | 1 | 3D SPGR | Volumetric and Cortical Thickening Analysis (FreeSurfer) |
| Ghisays et al. (2019) | - | - | - | - | - | Volumetric Analysis (Statistical Parametric Mapping) |
| Cotta Ramusino et al. (2019) | 1.5  1.5 | 20/2  21/20 | 30^0^  10^0^ | 1.3  2.0 | - | Visual Rating |
| Lyall et al. (2019) | - | - | - | - | - | Segmentation (FAST) |

TR, repetition time; TE, echo time; TI, inversion time; FA, flip angle; ST, slice thickness; SPGR: spoiled gradient recalled echo; IR-SPGR: inversion recovery-SPGR; FFE: fast field echo; 3D MP-RAGE: three dimensional magnetization-prepared rapid gradient-echo; FLAIR: fluid-attenuated inversion recovery; SE: spin echo; MDEFT: modified driven equilibrium Fourier transform; TFE: turbo field echo; FSL: FMRIB Software library; MDD: major depressive disorder

Table 2. Evidence table of studies included in qualitative synthesis

| Authors | Patient Group  (age in years) | Control group  (age in years) | APOE ε4 and Brain Structural Alterations | Remarks |
| --- | --- | --- | --- | --- |
| Cacciaglia et al. (2018) | None | 213 ♂and 320 ♀ (57.58 ± 7.43 years) | APOE ε4 carriers showed a reduced GMv in the right posterior hippocampus, but increased volume in the right medial thalamus relative to controls.  APOE ε4 dose dependent GMv increase was found in the left middle occipital cortex and in the right superior frontal cortex.  Additive effect of APOE ε4 on GMv reduction in the right caudate nucleus, right precentral gyrus, and right cerebellar crus. | Participants stratified into non-APOE ε4 and APOE ε4 zygosity.  Cross-sectional study. |
| Groot et al. (2018) | 35 (mean age: > 60 years) MCI;  145 (mean age: > 60 years) AD | 72 (66.6 ± 7.5 years) | Increased temporoparietal volume in APOE ε2 carriers relative to APOE ε4 carriers. | Patients stratified into APOE ε4 zygosity.  Cross-sectional study. |
| Kirsebom^*^ et al. (2018) | 1. 8 ♀ and 10 ♂ (66.7 ± 6.8 years) SCD;  2. 12 ♀ and 8 ♂ (66.8 ± 7.4 years) MCI | 1. 10 ♀ and 10 ♂; APOE ε4 (62.8 ± 9.6 years)  2. 9 ♀ and 7 ♂; APOE ε4- (59.1 ± 8.5 years) | Lack of associations between CSF biomarkers or APOE ε4 carrier and hippocampal or amygdala volumetry. | Cross-sectional study |
| Koval et al. (2018) | 154 AD (> 60 years) | None | Association between increased number of APOE ϵ4 genes and faster pace of cortical atrophy in AD but not with an earlier atrophy onset. | No information on gender distribution.  Longitudinal study. |
| Wang et al. (2018) | None | 266 ♀ and 170 ♂ (70.3 ± 9.1 years) | APOE ϵ4 shows an interaction with neurodegeneration scores on MMSE decline.  At baseline, no interaction was found between APOE ϵ4 and baseline and neurodegeneration scores. | Participants stratified into presence or absence of APOE ϵ4.  Neurodegeneration score include MRI markers of reduced hippocampus, and reduced total GM.  Longitudinal study |
| Wang et al. (2019) | 41.9%♀ and 58.1%♂ (72.6 ± 7.4 years) aMCI;  43.8%♀ and 56.2% ♂ (74.7 ± 8.0) AD | 49.1%♀ and 50.9% ♂; (74.7 ± 5.6) | MCI and AD APOE ε4 carriers demonstrated reduced hippocampal volumes relative to non-carriers. | Cross-sectional study.  Data derived from ADNI |
| Mattsson et al. (2018) | 10 ♀ and 9 ♂; (70.1 ± 7.8 years) APOE ε4-;  28 ♀ and 18 ♂; (72.4 ± 6.8 years) APOE ε4+ | None | Cortical thinning more pronounced in medial and lateral parietal regions of non-APOE ε4 carriers compared to APOE ε4 carriers. | Participants were classified as MCI due to AD/AD-like dementia.  PET included.  Cross-sectional study. |
| Moon et al. (2018) | 20 ♀ and 15 ♂ (63.81 ± 3.19 years) MCI;  8 ♀ and 7 ♂ (63.64 ± 3.01 years) AD | None | Increased atrophy rate of the left hippocampus in the MCI and AD APOE ε4 carriers compared to the right hippocampus of both groups. | Longitudinal study |
| Rane et al. (2018) | 14 ♀ and 47 ♂ (75.0±6.0); AD-MCI  11 ♀ and 22 ♂ (70.0±4.0); PD-MCI | 33 ♀ and 76 ♂ (75.0±5.0); CN  10 ♀ and 18 ♂ (70.0±4.0); CN | CN APOE ε4 carriers demonstrated decreased cortical thickness in the parahippocampal gyrus with increasing age.  Aβ negative APOE ε4 carriers demonstrated increased cortical thickness in the inferior parietal, middle temporal gyrus, and precuneus regions.  aMCI APOE ε4 carriers demonstrated reduced cortical thickness in the middle temporal gyrus, inferior parietal region, and the precuneus compared to non-carriers. | Atrophy of the precuneus, appearing similar in APOE ε4 carriers with AD and PD-MCI..  Cross-sectional study.  Data derived from ADNI |
| Sundermann et al. (2018) | MCI (306 ♀ and 324 ♂);  AD (135 ♀ and 178 ♂) | 335 ♀ and 367 ♂ | In the control, APOE ε4 allele was associated with reduced hippocampal volume ratio and hypometabolism in males but not in females.  In MCI, APOE ε4 was associated with reduced hippocampal volume ratio and hypometabolism and increased Aβ burden.  In AD, APOE ε4 carriers exhibited reduced hippocampal volume ratio relative to non-carriers. | PET included.  Cross-sectional study.  Data derived from ADNI.  Mean age: > 70 years |
| Dong et al. (2019) | None | 11♂ and 25♀ (57.2 ± 3.8 years) HT;  9♂ and 28 ♀ (58.4 ± 6.8) HM;  15♂ and 29♀ (58.6 ± 7.2 years). | Evidence of APOE ε4 gene-dose effects on the left hippocampal morphology (shape and atrophy). | Participants stratified according to the presence or absence of APOE ε4.  Cross-sectional study |
| Barboriak et al. (2000) | None | 92 (55 – 85 years) | APOE ε4 effect noted on hippocampal sulcal cavity. | Cross-sectional study |
| Lehtovirta et al. (1995) | 14♂ and 12♀ (mean age: > 65 years) AD | 6♂ and 10♀ (70.2 + 4.7 years) | AD APOE ε4 carriers exhibited reduced left frontal lobe, right hippocampal, and right amygdala volumes compared to controls. | Cross-sectional study |
| Du et al. (2006) | None | 42 (58 – 87 years) | No effect of APOE ε4 on atrophy rates of entorhinal cortex and hippocampus. | Longitudinal study |
| Barber et al. (2000) | 19♂ and 8♀ (75.9 ± 6 7 years) dementia with lewy bodies (DLB);  9♂ and 16♀ (77.2 ± 6 5 years) AD;  15♂ and 9♀ (76.9 ± 6 7 years) Vascular dementia (VaD) | 14♂ and 12♀ (76.2 ± 6 5 years) | No effect of APOE ε4 on volumetric measures of whole-brain, frontal, temporal lobe, hippocampal and amygdala. | Cross-sectional study |
| Boccardi et al. (2004) | 3♂ and 15♀ (76 ± 5 years) AD;  6♂ and 2♀ (62 ± 5 years) FTD | 9♂ and 17♀ (69 ± 9 years) | FTD APOE ε4 carriers had right ventral striatal atrophy.  AD APOE ε4 carriers had greater atrophy of the amygdalae and hippocampi head. | Participants stratified according to the presence or absence of APOE ε4.  Cross-sectional study |
| Reiman et al. (1998) | None | 33 (50 – 62 years) | No association found between APOE ε4 and hippocampal volume. | Cross-sectional study |
| Schmidt et al. (2008) | None | 106♂ and 108♀ (60.5 ± 6.0 years) | No association found between APOE ε4 and hippocampal and parahippocampal volumes, and sulcal widening. | Cross-sectional study |
| Basso et al. (2006) | 26♂ and 29♀ (mean age: > 70 years) AD | 22♂ and 20♀ (73.2 ± 6.7 years) | APOE ε4 effect observed on amygdala volume | Patients with AD stratified according to the presence or absence of APOE ε4.  Cross-sectional study |
| Espeseth et al. (2006) | None | 110 (53–64 years) middle-aged;  120 (65–75 years) older-adults | No association between APOE ε4 and total cortical volume. | Participants stratified according to the presence or absence of APOE ε4.  Cross-sectional study |
| Tanaka et al. (1998) | 34 (mean age: > 80 years) | 22 ( 82.0 ± 7.7 years) | AD patients with the APOE ε4 demonstrated severe  atrophy of the hippocampus and amygdala (with dilatation of the lateral ventricular temporal horn) relative to non-carriers. | Patients with AD stratified according to the presence or absence of APOE ε4  Cross-sectional study |
| Bigler et al. (2002) | 31♂ and 54♀ (81.04 ± 14.23 years) AD;  16♂ and 14♀ (84.09 ± 6.79 years) mild ambiguous/MCI  27♂ and 32♀ (82.22 ± 7.16 years) “mixed neuropsychiatric individuals” | 9♂ and 11♀ (76.88 ± 6.48 years) | No evidence of APOE ϵ4 effect on leftward asymmetry of hippocampus and parahippocampal gyrus. | Left asymmetry implies right > left in volumetric measures.  Cross-sectional study |
| Barber et al. (1999) | 9♂ and 16♀ (77.8 ± 4.4) AD  14♂ and 10♀ (76.9 ± 6.7 years) VaD  14♂ and 8♀ (77.2 ± 6.3 years) DLB | None | No evidence of an association between APOE ϵ4 and medial temporal lobe atrophy. | Cross-sectional study |
| Carmelli et al. (2000) | None | 308 (72.5 ± 3.0 years) non-APOE ε4 carriers  82 (71.9 ± 2.8 years) APOE ε4 carriers | APOE ε4 carriers exhibited reduced total intracranial (whole brain) volume and increased CSF volume. | No information on gender distribution.  Longitudinal study |
| Pennanen et al. (2006) | 51 MCI (mean age: > 70 years) | 13♂ and 19♀ (73 ± 4 years) | Homozygous APOE ε4 carriers showed more atrophy of the bilateral amygdala, right parahippocampal gyrus, left medial dorsal thalamic nucleus, and the bilateral temporoparietal regions relative to non-carriers. | Cross-sectional study |
| Morra et al. (2009) | 160♂ and 85♀ (74.99 ± 7.17 years) MCI  49♂ and 48♀ (75.77 ± 7.33 years) AD | 75♂ and 73♀ (75.92 ± 4.90 years) | Correlation found between APOE ε4 and the rate of decreased hippocampal volume in both AD and controls.  APOE ε4 carriers showed faster loss of hippocampal tissue compared to non-carriers. | Longitudinal study  Data derived from ADNI. |
| Goltermann et al. (2019) | 24 MDD | 51.6%♀ (34.47 ± 13.48 years) | No evidence of APOE ε4 effect on hippocampal volume.  Reduced mean total intracranial volume (whole brain) in APOE ε4 homozygotes. | Participants stratified into mild depressive disorder (n = 62) homozygous APOE ε4 and non-APOE ε4 carriers.  Cross-sectional study. |
| Konishi et al. (2018) | None | 28♂ and 38♀ (66.1 ± 4.5 years) | APOE ɛ4 carriers demonstrated less grey matter in the entorhinal cortex relative to the non-carriers. | Multimodal study with fMRI.  Cross-sectional study. |
| Kelly et al. (2018) | None | 15 declining APOE ε4 carriers  22 declining non-APOE ε4  11 APOE ε4 carriers (cognitively stable) | APOE ε4 declining group exhibited greater atrophy in the bilateral cortical GMvs and bilateral hippocampi relative to non-carriers. | Longitudinal study; Age range: 65–85 years. |
| Alemany et al. (2018) | None | 212♂ and 172♀(8.49 ± 0.84 years) APOE ϵ4 carriers  673♂ and 610♀ (8.53 ± 0.88 years) non-APOE ϵ4 carriers | APOE ε4 status did not show an association with volumetric sizes of the basal ganglia including caudate, putamen, and globus pallidum. | Cross-sectional study |
| Tosun et al. (2010) | 73♂ and 46♀ (74 ± 7.6 years) MCI  29♂ and 25♀ (74 ± 8.0 years) AD | 37♂ and 40♀ (75 ± 5.0 years) | In MCI group, CSF biomarker levels (i.e., reduced Aβ42, increased p-tau181p and t-tau) and APOE ε4 exhibited independent association with brain tissue volume loss in bilateral hippocampus, bilateral temporal lobe (i.e, middle temporal, inferior temporal, temporal pole, fusiform), right amygdala, right lingual, left entorhinal, left isthmus cingulate, left parahippocampal, and left precuneus cortices.  In MCI group, reduced CSF Aβ42 and APOE ε4 were observed to show association with increased rates of cortical thinning in temporoparietal cortex, precuneus and PCC.  In MCI group, both CSF tau and APOE ε4 showed association with increased rates of cortical thinning in the temporal pole, entorhinal cortex, and precuneus.  In the AD group, reduced CSF Aβ and increased t-tau levels and the presence of APOE ε4 were associated with increased rates of brain tissue volume loss in the bilateral caudate.  In AD, APOE ε4 showed an association with increased rate of volume loss in caudate, seen to be independent of t-tau level. | Longitudinal study  Data derived from ADNI. |
| Hua et al. (2008) | 330 (74.8 ± 7.5 years) MCI  165 (75.6 ± 7.6 years) AD | 181 (75.9 ± 5.1 years) | Association was found between one copy of APOE ε4 and increased CSF expansion in the Sylvian fissures, between inferior frontal and superior temporal lobes.  Association was found between increased atrophy of the hippocampal and temporal lobe and carriership of an additional APOE ε4 allele. | Cross-sectional study  Data derived from ADNI. |
| Filippini et al. (2009) | None | 59♂ and 101♀ (55 ± 11 years) non-APOE ϵ4 carriers  16♂ and 35♀ (53 ± 14 years) APOE ϵ4 carriers | Faster age‐related decline and decreased volumes of the corpus callosum and its subregions connected to the prefrontal, premotor, motor, sensorial, posterior parietal, temporal, and occipital cortices  is faster in APOE ϵ4 carriers than noncarriers | Cross-sectional study |
| Jak et al. (2007) | None | 21♂ and 26♀ (77.6 ± 6.7 years) non-APOE ε4  12♂ and 10♀ (74.9 ± 7.2 years) APOE ε4 carriers | APOE ε4 carriers showed increased rate of hippocampal atrophy relative to non-carriers. | Longitudinal study |
| Doody et al. (2000) | 104 AD (mean age: > 60 years) | None | No association between APOE ε4 and atrophy of the frontal, parietal, temporal, occipital, basal ganglion, and cerebellum. | Cross-sectional study |
| Geroldi et al. (2000) | 28 AD | 10♂ and 20♀ (69 ± 8 years) | Increased APOE ε4 gene dose on hippocampal atrophy (Right < left) in AD compared to controls  No effect of APOE ε4 on temporal lobe and entorhinal cortex. | AD patients stratified into presence or absence of APOE ε4; Cross-sectional study. |
| Mishra et al. (2018) | None | 189♂ and 308♀ (66.8 ± 10.0 years) | Higher rate of subcortical atrophy of the hippocampus, amygdala, putamen, and accumbens in APOE ɛ4 carriers compared to non-carriers. | Longitudinal study; PET data included. |
| Bussy et al. (2019) | None | 46♂ and 72♀ (37.18 ± 10.54 years) non-APOE ε4 carriers  18♂ and 26♀ (43.05 ± 11.38 years) APOE ε4 carriers | No differences between APOE ε4 carriers and non-carriers on precuneal and hippocampal volumes. | Cross-sectional study |
| Taylor et al. (2014) | None | 22♂ and 9♀ (62.2 ± 6.3 years) non-APOE ε4 carriers  23♂ and 2♀ (59.4 ± 5.7 years) APOE ε4 carriers | APOE ε4 carriers showed reduced hippocampal volumes relative to non-carriers. | Longitudinal study |
| Kerchner et al. (2014) | 9♂ and 5♀ (73.2 ± 5.3) aMCI  5♂ and 6♀ (69.5 ± 9.3) AD | 7♂ and 7♀ (68.8 ± 4.7) | Association of APOE ε4 dose-dependent with increased thinning of the hippocampal CA1 apical neuropil, or stratum radiatum/stratum lacunosum-moleculare. | Cross-sectional study |
| Fennema-Notestine et al. (2011) | None | 482 (51 – 59) | APOE ε4 carriers exhibited thinner cortex in bilateral superior frontal, left rostral midfrontal, and right caudal midfrontal regions.  No effect of APOE ε4 on the hippocampus, entorhinal, parahippocampal, lateral temporal (inferior, middle, and superior temporal), and frontal (caudal and rostral middle; superior; inferior; orbitofrontal) cortex. | Participants stratified according to the presence or absence of APOE ε4; Longitudinal study; Only men were represented. |
| Andrawis et al. (2012) | 243 MCI  96 AD | 145 | APOE ε4 carriers demonstrated reduced hippocampal volume, and APOE ε4 was found to be associated with reduced hippocampal volume at follow-up compared with non-carriers. | Participants stratified according to the presence or absence of maternal/family history of dementia.  Longitudinal study.  Mean age: > 70 years  Data derived from ADNI. |
| Chang et al. (2016) | None | 1187 (3 to 20 years) | APOE ε4 carriers demonstrated the smallest hippocampi, and entorhinal cortical thinning of the left inferior parietal gyrus and right superior parietal gyrus.  APOE ε4 carriers demonstrated the largest medial orbitofrontal cortical areas. | Cross-sectional study |
| Honea et al. (2009) | None | 24♂ and 15♀ (73.7 ± 6.2 years) non-ε4 carriers  6♂ and 8♀ (72.7 ± 6.2 years) | APOE ε4 allele carriers showed atrophy of the left anterior hippocampus and amygdala relative to non-carriers. | Cross-sectional study |
| Chang et al. (2014) | 104 AD | 123 | No APOE ε4 effects on volumetric measures of bilateral hippocampal (including the dentate gyrus, CA fields, subiculum/parasubiculum and fimbria), and cortical thickness of frontal, parietal lobe cortical regions, temporal, and bilateral cingulate cortical regions in the AD groups. | Patient group and control group classified as young-old and very-old with or without APOE ε4; Longitudinal study.  Data derived from ADNI.  Mean age: > age > 70 years |
| Li et al. (2016) | 353 (mean age: 75.06 years) MCI  160 (mean age: 74.88 years) AD | 211 (mean age: 76.41 years) | APOE ε4 carriers demonstrated morphological deformation of the hippocampus relative to non-carriers in the entire cohort.  APOE ε4 carriers demonstrated increased left hippocampal atrophy relative to the right hippocampus. | Participants stratified according to the number of samples recruited in 6-, 12- and 24-months, and the presence or absence of APOE ε4. Longitudinal study; Sample size included were that of 6 months.  Data derived from ADNI. |
| Okonkwo et al. (2010) | 166♂ and 92♀ (74.48 ± 7.25 years) MCI  56♂ and 50♀ (75.11 ± 7.35 years) AD | 84♂ and 80♀ (76.03 ± 5.22 years) | APOE ε4 carriers demonstrated increased whole brain atrophy in MCI. | Cross-sectional study  Data derived from ADNI. |
| Tosun et al. (2011) | 73♂ and 46♀ (74 ± 7.6 years) MCI;  29♂ and 25♀ (74 ± 8.0 years) AD | 37♂ and 40♀ (75 ± 5.0 years) | APOE ε4 was found to be associated with higher rates of cortical thinning in entorhinal cortex, temporoparietal cortex, temporal pole, PCC, and precuneus in MCI individuals. | Longitudinal study  Data derived from ADNI. |
| Tang et al. (2015) | 276 MCI  129 AD | 135 | Atrophy of the hippocampi and bilateral amygdala in APOE ε4 carriers relative to the non-carriers, specifically of the young-old participants.  Reduced hippocampal volume in APOE ε4 carriers relative to the non-carriers in those with AD.  Reduced surface area (or shape atrophy) of the right hippocampus, and bilateral amygdala in AD-young APOE ε4 carriers relative to AD-young non-carriers. | Patient group and control group classified as young-old (< 75 years) and very-old (> 80 years) with or without APOE ε4.  Longitudinal study.  Data derived from ADNI. |
| Burggren et al. (2008) | None | 7♂ and 7♀ (57.7 ± 9.6 years) APOE ε4 carriers  8♂ and 8♀ (57.3 ± 7.8 years) non-APOE ε4 carriers | APOE ε4 carriers demonstrated lower cortical thickness in the entorhinal cortex and subiculum relative to non-carriers. | Cross-sectional study |
| Reiter et al. (2017) | None | 7♂ and 23♀ (73.94 ± 5.38) non-APOE ε4 carriers  14♂ and 28♀ (73.02 ± 4.98) APOE ε4 carriers | APOE ε4 carriers exhibited greater rate of atrophy in total GM, right hippocampal subfields (CA 1-4, dentate gyrus, presubiculum/subiculum and fimbria), bilateral hippocampi, parahippocampal gyrus, bilateral lingual gyri, and right lateral orbitofrontal cortex relative to the non-carriers. | Longitudinal study |
| Lampert et al. (2014) | None | 184 (mean: 79.9 years) | APOE ε4 effect observed on atrophy of hippocampus, amygdala, entorhinal cortex and cerebral cortex. | Longitudinal study  Data derived from ADNI. |
| Bender and Raz (2012) | None | 22♂ and 50♀ (19 to 77 years) | Older APOE ε4 with higher blood pressure exhibited reduced lateral prefrontal cortex volumes. | Participants stratified according to the presence or absence of APOE ε4, Cross-sectional study. |
| Mueller and Weiner (2009) | 12♂ and 6♀ (69.1 ± 9.6 years) AD;  14♂ and 6♀ (73.5 ± 7.1 years) aMCI | 55♂ and 64♀ (53.4 ± 17.2 years) | Association between AD APOE ε4 and volume loss in CA3-dentate gyrus relative to non-APOE ε4 carriers in HCs and AD. | Participants stratified according to the presence or absence of APOE ε4; Cross-sectional study. |
| Mueller et al. (2008) | (67.5 ± 9.3 years) AD | 81 (60.8 ± 13.6 years) | Reduced CA3&DG in AD with APOE ε4 relative to AD without APOE ε4.  APOE ε4 effect observed on CA3-dentate gyrus demonstrating reduced volumes only in the entire control population, older controls, a subgroup of older subjects, and AD subjects. | Control group stratified as young and old age adults with or without APOE ε4, Cross-sectional study. |
| Donix et al. (2010) | None | 4♂ and 12♀ (60.1±7.1 years) non-APOE ε4 carriers  6♂ and 10♀ (61.7±11.5 years) APOE ε4 carriers | Lower cortical thickening in the subiculum and entorhinal cortex of APOE ε4 carriers relative to non-carriers.  Reduced cortical thickening across the medial temporal lobe sub-regions (cornu ammonis fields 1, 2, and 3, dentate gyrus, subiculum, entorhinal cortex, perirhinal cortex, parahippocampal cortex, and fusiform gyrus) combined in APOE ε4 carriers relative to non-carriers. | Longitudinal study |
| Ferencz et al. (2013) | None | 424 (60 – 87 years) | No evidence of APOE effect on hippocampal volume. | Participants stratified according to TOMM40 genotype and presence or absence of APOE ε4 allele; Cross-sectional study. |
| Spampinato et al. (2011) | 48 MCI (mean age: > 70 years)  47 MCI-AD (mean age: > 70 years) | None | APOE ε4 carriers exhibited GMv loss of the temporal lobe, hippocampi, parietal lobe, right caudate nucleus, and insulae in MCI to AD group.  APOE ε4 carriers who were MCI stable exhibited lower GMv of the temporal lobes and bilateral insular. | Longitudinal study  Data derived from ADNI. |
| Ma et al. (2016) | None | 836 (65.2 ± 7.5 years) | APOE ε4 carriers demonstrated reduced GM of the right fusiform gyrus, right inferior temporal gyrus, left middle and superior frontal gyrus, right angular gyrus, and left cingulate gyrus and precuneus relative to non-carriers. | Participants stratified into 4 groups according to rs405509 T allele and APOE ε4 allele. Cross-sectional study. |
| Lu et al. (2011) | None | 5♂ and 6♀ (67.0 ± 5.2 years) non-APOE ε4 carriers  7♂ and 9♀ (65.0 ± 4.5 years) APOE ε4 carriers | APOE ε4 exhibited greater annual atrophy rates in the temporal lobes and hippocampus (right > left). | Longitudinal study |
| Dean et al. (2014) | None | 60 APOE ε4 carriers  102 non-APOE ε4 carriers | APOE ε4 carriers exhibited GMv reduction of PCC, medial cingulate cortex, precuneus, lateral temporal, and medial occipitotemporal regions relative to non-carriers. | All participants were infants with age range between 2- to 25-month-old, cross-sectional study. |
| Wolk et al. (2010) | 38♂ and 29♀ (74.9 ± 9.2 years) APOE ε4 carriers  13♂ and 11♀ (74.3 ± 7.3 years) non-APOE ε4 carriers | None | APOE ε4 carriers demonstrated reduced hippocampal volume relative to non-carriers.  APOE ε4 carriers demonstrated reduced cortical thickness of the superior parietal lobule, precuneus, angular gyrus, and superior frontal gyrus relative to non-carriers.  APOE ε4 carriers demonstrated more cortical thinning of the superior and midline parietal and dorsolateral frontal regions, in addition to medial temporal, caudal temporal, lateral temporal regions, and occipital regions relative to the non-carriers.  Non-carriers exhibited greater frontoparietal atrophy. | All participants were AD patients, Cross-sectional data.  Data derived from ADNI. |
| Soldan et al. (2015) | None | 148♂ and 201♀ (57.3 ± 10.4 years) | No associations between APOEε4 and rate of atrophy of entorhinal cortex and hippocampus.  Left amygdala showed an interaction between atrophy rate and APOE ε4 genotype in association with symptom onset. |  |
| Fan et al. (2010) | None | 75♂ and 84♀ (mean age: >70 years) | Increased cortical thickness of the left superior temporal and left dorsolateral prefrontal region in APOE ε2 carriers relative to APOE ε4 carriers. | Participants stratified according to APOE ɛ4 genotypes.  Data derived from ADNI. |
| Banks et al. (2017) | None | 54 (30.1 ± 9.3 years) APOE ε4 carriers  139 (31.1 ± 9.4 years) non-APOE ε4 carriers | No impact of APOE ε4 on volumes of bilateral hippocampus, bilateral thalamus, and bilateral caudate. | (Professional fighters, cross-sectional study, no information on gender distribution) |
| Walsh et al. (2013) | 88 AD (mean age: > 70 years) | None | No association found between the APOE ε4 dose and hippocampal volume in this cohort. | Cross-sectional study |
| Li et al. (2017) | 165♂ and 132♀ (71.53 ± 7.43 years) Early MCI  111♂ and 85♀ (73.83 ± 8.06 years) Late MCI  94♂ and 86♀ (74.94 ± 7.81 years) AD | 123♂ and 128♀ (75.47 ± 6.54) | Among CN and early MCI participants, there was association between APOE ε4 allele and increased cortical thickening in the entorhinal cortex, parahippocampal gyrus, inferior temporal gyrus, and temporal pole relative to the non-carriers.  Late MCI and AD participants who are APOE ε4 carriers exhibited reduced cortical thickness relative to the non-carriers. | Multimodal study with PET included.  Cross-sectional study.  Data derived from ADNI. |
| Chen et al. (2012) | None | 27 APOEε4 homozygotes  36 APOE ε4 heterozygotes  67 non-APOE ε4 carriers | Increased APOE-ε4 dose found to be associated with reduced GMv in inferior frontal, parietal, anterior cingulate, temporal, parahippocampus, and hippocampus. | Multimodal study with FDG-PET included, longitudinal cohort study.  Mean age: > 55 years |
| Hostage et al. (2014) | 237 (mean age= 79.9 years) MCI | None | Association found between APOE ε4 allele and accelerated rates of atrophy of the amygdala, cerebral cortex, entorhinal cortex, fusiform gyrus, hippocampus, inferior parietal cortex, inferior temporal cortex, middle temporal cortex, parahippocampal cortex, precuneus cortex, superior parietal cortex, temporal pole cortex, and transverse temporal cortex. | Longitudinal study  Data derived from ADNI. |
| Donix et al. (2013) | 28 (mean age: > 72 years) AD | 26 (mean age: > 55 years) middle-age individuals;  23 ( mean age: > 65 years) older individuals | APOE ε4 carriers exhibited thinner entorhinal cortex of the left hemisphere relative to the right hemisphere. | Cross-sectional study |
| Shi et al. (2014) | 167 (mean age: > 70 years) AD;  354 (mean age: > 70 years) MCI | 204 (mean age: > 70 years) | Accelerated hippocampal atrophy (left > right) more salient in APOE ε4 homozygotes relative to heterozygotes. | Cross-sectional study  Data derived from ADNI. |
| Hafsteinsdottir et al. 2012 | None | 1280♂ and 1845♀ (76.3 ± 5.4years) non-APOE ε4 carriers  505♂ and 673♀ (75.7 ± 5.3 years) APOE ε4 carriers | APOE ε4 carriers demonstrated reduced GMvs compared to non-carriers. | Longitudinal study |
| Novellino et al. (2019) | 95 MCI (mean age > 70 years) | 15♂ and 34♀ (71.6 ± 4.26) | Increased atrophy of GM in hippocampus, parahippocampal gyrus, and thalamus in MCI homozygous-APOE ε4 relative to MCI heterozygous-APOE ε4 and MCI non-APOE ε4 controls.  More marked GM atrophy of bilateral middle frontal gyrus in MCI non-APOE ε4 relative to MCI heterozygous-APOE ε4 and MCI homozygous-APOE ε4 groups. | Cross-sectional study |
| Juottonen et al. (1998) | 5♂ and 6♀ (69.1 ± 7.1 years) APOE ε4 –  10♂ and 6♀ (70.4 ± 9.9 years) APOE ε4 + | 11♂ and 20♀ (72.2 ± 3.9 years) | APOE ε4 is associated with atrophy of the entorhinal cortex in early AD. | Cross-sectional study  Patient group are classified as AD. |
| Wilhelm et al. (2008) | None | 23♂ and 9♀ (45.97 ± 7.5 years) non-APOE ε4 allele carriers  11♂ and 9♀ (47.55 ± 11.1 years) APOE ε4 allele carriers | No effect of APOE ε4 on reduced hippocampal volume. | Cross-sectional study |
| Cherbuin et al. (2008) | None | 177♂ and 154♀ (62.6 ± 1.4 years) | No association between APOE ε4 allele and atrophy of the hippocampus and amygdala | Longitudinal study |
| Biffi et al. (2010) | 74♂ and 143♀ (75.3 ± 7.4 years) MCI-nc  55♂ and 85♀ (74.6 ± 6.8) years) MCI-c  81♂ and 87♀ (75.5 ± 7.7 years) AD | 97♂ and 118♀ (75.9 ± 5.5 years) | Association found between APOE ε4 allele and reduced volumes of the hippocampus and amygdala, reduced thickness of entorhinal cortex, parahippocampal gyrus cortex, and temporal lobe cortex. | Longitudinal study  Data derived from ADNI. |
| Ystad et al. (2009) | None | 170 (46 – 77 years) | No associations between hippocampal volumes and APOE ε4 allele | Cross-sectional study |
| Schuff et al. (2009) | 140♂ and 86♀ (75.0 ± 7.1 years) MCI  51♂ and 45♀ (75.8 ± 6.6 years) AD | 66♂ and 61♀ (76.3 ± 5.1 years) | Association found between increased rates of hippocampal atrophy and APOE ε4 allele in AD | Longitudinal study  Data derived from ADNI. |
| Stewart et al. (2011) |  | 510♂ and 826♀ (72.0 ± 4.0 years) | Associations between reduced hippocampal volume and subjective memory impairment at follow-up, found to be stronger in APOE ε4 allele | Longitudinal study |
| Protas et al. (2013) | None | 28♂ and 48♀ (56.5 ± 4.7 years)  15♂ and 27♀ (55.9 ± 4.0 years) HT APOE ε4  10♂ and 21♀ (55.5 ± 5.1 years) HM APOE ε4 | Association found between APOE ε4 and reduced hippocampal volume | Cross-sectional study |
| Hoogendam et al. (2012) | None | 1806♂ and 2156♀ (60.1 ± 8.50 years) | No association found between APOE ε4 and cerebral or cerebellar volume. | Cross-sectional study |
| Geroldi et al. (1999) | 28 AD | 10♂ and 20♀ (69 ± 8 years) | Progressive reduction of volumes of the temporal lobe regions volumes (i.e. entorhinal cortex and hippocampus) from controls to AD patients with increasing APOE ε4 gene-dose.  Increased frontal lobe volume with increasing APOE ε4 gene dose in AD patients | Cross-sectional study |
| Lehtovirta et al. (1996) | 30♂ and 28♀; AD | 14♂ and 20♀ | AD homozygous APOE ε4 allele carriers demonstrated atrophy of the hippocampus and amygdala relative to control group.  No association between ε4 allele and frontal lobe. | Cross-sectional study  Mean age: > 60 years |
| O’Dwyer et al. (2012) | None | 13♂ and 9♀ (26.86 ± 5.28 years) APOE ε4 carriers  13♂ and 9♀ (26.73 ± 4.00 years) non-APOE ε4 carriers | Lower hippocampal volume (right < left) in healthy young APOE ε4 carriers compared to non-carriers | Cross-sectional study |
| Sabuncu et al. (2012) | 58♂ and 42♀ ( 75.1 ± 7.8 years) AD | 54♂ and 50♀ (75.9 ± 5.1 years) | No association found between APOE ε4 allele and cortical thickness of entorhinal cortex, temporopolar cortex, lateral temporal cortex, inferior parietal cortex and sulcus, PCC, and inferior frontal cortex | Cross-sectional study  Data derived from ADNI. |
| Bunce et al. (2012) | None | 314 (44 – 48 years)  313 (64 – 68 years) | No association between APOE ε4 and bilateral entorhinal cortex, bilateral middle and superior temporal region, and bilateral inferior and temporal poles. | Longitudinal study |
| Hostage et al. (2013) | 321 mild MCI  143 AD | 198 | Evidence of significant dose-dependent interactions between APOE ε4 and the diagnoses of MCI and AD resulting in reduced hippocampal volumes in both groups (AD < MCI) relative to the lack of effect of APOE ε4 on hippocampal volume of HC group. | Cross-sectional study  Data derived from ADNI.  Mean age: > 70 years |
| DiBattista et al. (2014) | None | 33♂ and 24♀ (21.8 ± 4.0 years) | APOE ε4 carriers demonstrated increased right entorhinal cortex volume relative to non-carriers | Cross-sectional study |
| Manning et al. (2014) | 81♂ and 67♀ (75.0 ± 7.6 years) AD  307 MCI | 90♂ and 77♀ (76.0 ± 5.1 years) | Increased hippocampal atrophy rates in APOE ε4 carrier AD, MCI-progressors and MCI-stable relative to APOE ε4 non-carriers. | Longitudinal study  Data derived from ADNI. |
| Khan et al. (2014) | None | 2842 | No evidence of dose-dependent effect of APOE ε4 alleles or APOE ε2 alleles on either hippocampal volume and hippocampal asymmetry | Cross-sectional study  Mean age: 14.44 ± 0.41 years |
| Holland et al. (2013) | 273 MCI  110 MCI-converter  105 AD | 188 | Significant effects of APOE ε4 found on structures of the medial temporal lobe (i.e. entorhinal cortex and hippocampus), inferior parietal cortex, in addition to additive effect of APOE ε4 on annual atrophy rate | Longitudinal study  Data derived from ADNI.  Mean age: > 70 years |
| Lyall et al. (2013) | None | 343♂ and 312♀ | APOE ε4 allele showed no effect on hippocampal volumes. | Longitudinal study  Mean age: 55-76 years |
| Morgen et al. (2015) | 57♂ and 63♀ ( 70.4 ± 6.4 years) AD APOE ε4 carriers  28♂ and 35♀ (70.4 ± 8.7 years) AD APOE ε4 non-carriers |  | No association was found between APOE ε4 allele and decreased right hippocampus volume.  No association was found between non-carriers status of APOE ε4 and reduced volume of the right superior frontal gyrus relative to carriers. | Longitudinal study |
| Risacher et al. (2015) | 104 SMC  305 early MCI | 185 | Reduced hippocampal volume in early MCI APOE ε4 carriers and noncarriers relative to CN APOE ε4 noncarriers and SMCI APOE ε4 carriers. Reduced hippocampal volume was found in early MCI noncarriers of APOE ε4 relative to CN APOE ε4 carriers. | Cross-sectional study  Data derived from ADNI.  Mean age: > 70 years |
| Yokoyama et al. (2015) | None | 142 (39 – 83 years) | Reduced left cuneal volume found to be associated with APOE ε4. | Longitudinal study |
| Sampedro et al. (2015) | None | 90♂ and 78♀ (73.4 ± 6.02 years) MRI data  166♂ and 162♀ (74.5 ± 5.57 years) FDG-PET data  136♂ and 138♀ (74.4 ± 5.97 years) CSF data | Relative to the non-carriers, male APOE ε4 carriers exhibited increased cortical thickness in the left hemisphere, dorsolateral frontal region, temporoparietal, occipital and precuneus regions; and in the right hemisphere in the occipital and parietal regions.  APOE-by-gender interaction in the dorsolateral frontal and temporoparietal regions. | Cross-sectional study  Data derived from ADNI. |
| Khan et al. (2017) | 365 AD  522 MCI | 2,281 | Reduced hippocampal volume was found in APOE ϵ4 carriers relative to APOE ϵ3 carriers; APOE ϵ3 carriers and APOE ϵ2 carriers exhibited intermediate volumes and the largest volumes respectively.  AD and MCI APOE ϵ4 carriers demonstrated more reduced hippocampal volumes relative to APOE ϵ2 carriers with the highest hippocampal volumes.  Participants with both Aβ+ and APOE ϵ4 showed the lowest hippocampal volumes relative to Aβ- APOE ϵ4 carriers. | Cross-sectional study  Data derived from ADNI and other cohort studies (AddNeuroMed, AIBL, BRC-AD, SNAC-K, IMAGEN)  ADNI (age: 55-70 years); AIBL (at least 60 years); BRC-AD (mean age:> 70 years); SNAC-K (mean age: > 60 years); IMAGEN: (mean age: 14.4 years). |
| Falahati et al. (2017) | 15♂ and 60♀ (74.5 ± 7.4 years) MCI-stable  27♂ and 43♀ (74.3 ± 6.9 years) MCI-progressed  95♂ and 100♀ (75.5 ± 7.5 years) AD | 111♂ and 117♀ (75.9 ± 5.0 years) | Higher average severity index (SI) and a faster increase of SI of mediotemporal region in APOE ε4 positive MCI relative to APOE ε4 negative MCI. | Longitudinal study  Data derived from ADNI. |
| Rogne et al. (2016) | 14♂ and 11♀ (70.0 ± 9.1 years) SMC  18♂ and 115♀ (74.5 ± 7.5 years) MCI | 36♂ and 22♀ (70.6 ± 6.7 years) | Association was found between APOE ε4 and reduced hippocampal volume. | Longitudinal study |
| Konishi et al. (2016) | None | 64♂ and 56♀ (23.83 ± 4.32 years) Behavioural sample  19♂ and 18♀ (25.00 ± 4.26 years) MRI subsample | Increased GM of hippocampus, middle temporal gyrus, and precuneus in APOE ε2 allele relative to APOE ε3 and APOE ε4 carriers. | Cross-sectional study |
| Habes et al. (2016) | None | 1472 (22-90 years) | No evidence of an association between APOE ε4 and volumes of the lateral and medial frontal region, lateral temporal and hippocampus. | Longitudinal study |
| Fang et al. (2019) | 34♂ and 46♀ (69.2 ± 5.1y ears) MCI-AD  44♂ and 48♀ (72.8 ± 6.2 years) MCI-MCI | 46♂ and 44♀ (75.2 ± 6.8 years) | MCI-AD and MCI-MCI groups who were APOE ε4 carriers showed reduced hippocampal volume relative to that of the control group. | Longitudinal study |
| Lupton et al. (2016) | 280♂ and 284♀ (75.5 ± 7.1 years) AD  648♂ and 492♀ (75.8 ± 6.7 years) MCI | 746♂ and 1021♀ (75.3 ± 5.9 years) older adults  467♂ and 765♀ (22.9 ± 3.3 years) young adults | An association was found between APOE ε4 and lower volumes of the amygdala and hippocampus in AD and MCI but not in healthy older group. | Longitudinal study  Data derived from ADNI and other cohort studies. |
| Hobel et al. (2019) | 540♂ and 390♀ (55 – 90 years for males; 55 – 96 years for females) MCI | None | Significantly reduced left hippocampal volumes in females relative to males with 1 APOE ε4 allele.  Significantly reduced right hippocampal in females relative to males with 0, 1, or 2 APOE ε4 alleles  Significant reduction in bilateral amygdala for females with 0, 1, and 2 APOE ε4 alleles relative to men. | Cross-sectional study  Data derived from ADNI and other cohort studies |
| Schreiber et al. (2017) | 302 MCI  220 Late MCI | 125 ♂ and 131 ♀ (75.5 ± 6.7 years) | In MCI with suspected non–AD pathophysiology, association was found between APOE ε4 and reduced  GMv (left middle temporal region). | Cross-sectional multimodal (PET included) study.  Data derived from ADNI |
| Haller et al. (2017) | None | 43 (74.1 ± 3.8 years; 22 sCON and 21 dCON) APOE ε2  274 (74.1 ± 4.1 years; 132 sCON and 142 dCON) APOE ε3  65 (73.6 ± 4.1 years; 27 sCON and 38 dCON) APOE ε4. | Lower GM concentration of PCC and amygdala in APOE ε4 relative to APOE ε2 and APOE ε3.  Increased GM concentration of parietal lobe in APOE ε4 and APOE ε3 carriers. | Longitudinal study |
| Nao et al. (2017) | None | 12 ♂ and 20 ♀ (27.5 ± 5.0 years) APOE ε4 +  16 ♂ and 24 ♀ (28.1 ± 5.3 years) APOE ε4 – | APOE ε4 carriers demonstrated reduced GMv in the bilateral anterior and middle cingulate gyri. | Cross-sectional multimodal (task-based fMRI included) study. |
| Li et al. (2019) | None | 300 ♂ and 195 ♀ (70.14 ± 15.0 years) APOE ε4 +  1363 ♂ and 975 ♀ (70.9 ± 14.8 years) APOE ε4 – | AD APOE ε4 carriers demonstrate increased incidence of brain atrophy and severe brain atrophy. | Cross-sectional study |
| Hays et al. (2019) | None | 26 ♂ and 50 ♀ (72.98 ± 6.03 years) APOE ε4 +  13 ♂ and 28 ♀ (74.02 ± 6.62 years) APOE ε4 – | An interaction was found between APOE and right caudal anterior cingulate cortex (cACC) cortical thickness on memory performance, such that increased CT in the right cACC showed association with worse memory in APOE ε4 carriers relative to the non-carriers. | Longitudinal study |
| Herrmann et al. (2019) | None | 148 ♂ and 232 ♀ (74.2 ± 4.1 years) | No association between APOE ε4 and GMvs of hippocampus and amygdala. | Cross sectional study; PET included. |
| Foley et al. (2017) | None | 77 ♂ and 195 ♀ (24.8 ± 6.9 years) T1 data  34 ♂ and 53 ♀ (23.9 ± 4.4 years) Tractography data | Inclusion of APOE locus with polygenic risk scores (PRSs) demonstrated significant association between AD PRSs and reduced left hippocampal volume. | Cross-sectional study; DTI included |
| Ghisays et al. (2019) | None | 47–86 years (26 APOE ε4 HMs; 48 HTs; 90 NCs) | Cortical thickness and hippocampal volumes did not show any association with APOE ε4 in entire age range. | Cross-sectional study; PET included |
| Cotta Ramusino et al. (2019) | None | 20-84 years (50.2 ± 14.7 years) | No evidence of an association between APOE ε4 and medial temporal atrophy nor posterior atrophy severity. | Cross-sectional study |
| Lyall et al. (2019) | None | 2916 ♂ and 3124 ♀ (61.66 ± 7.03 years) APOE ε4 –  941 ♂ and 1206 ♀ (61.19 ± 7.04 years) APOE ε4 + | No evidence of an association between APOE ε4 and volumes of bilateral hippocampus, total GM. | Cross-sectional study |

AD: Alzheimer’s disease; GMvs: gray matter volumes; SCD: subjective cognitive decline; aMCI: amnestic mild cognitive impairment; EMCI: early mild cognitive impairment; LMCI: late mild cognitive impairment; PD: Parkinson’s disease; FTD: frontotemporal dementia; VaD: vascular dementia; DLB: dementia with Lewy bodies; DMN: default mode network; TIV: total intracranial volume; PPR: pulse pressure; KIBRA: KIdney and BRAin expressed protein; PRSs: polygenic risk scores; sCON: stable cognitive function; dCON: deteriorating cognitive function; ADNI: Alzheimer’s Disease Neuroimaging Initiative; AIBL: Australian, Imaging, Biomarkers and Lifestyles ; BRC-AD: Biomedical Research Centre for Dementia; SNAC-K: Swedish National study on aging and care in Kungscholmen; IMAGEN: Neuroimaging-Genetics

Table 3 Methodological quality of included studies

| Study | 1 | 2 | 3 | 4 | 5 | 6 | 7 | 8 | 9 | Total Score | LOE |
| --- | --- | --- | --- | --- | --- | --- | --- | --- | --- | --- | --- |
| Cacciaglia et al. (2018) | + | + | + | + | + | + | + | + | - | 8/9 | B |
| Groot et al. (2018) | + | + | - | + | + | + | + | + | - | 7/9 | B |
| Kirsebom et al. (2018) | + | + | - | + | + | + | + | + | - | 7/9 | B |
| Koval et al. (2018) | + | + | - | - | + | + | + | - | - | 5/9 | B |
| Wang et al. (2018) | + | - | + | + | + | + | + | + | + | 8/9 | B |
| Wang et al. (2019) | + | / | - | + | + | + | / | / | / | 4/9 | B |
| Mattsson et al. (2018) | + | / | + | - | + | + | + | - | + | 6/9 | B |
| Moon et al. (2018) | + | - | / | + | + | + | + | + | + | 7/9 | B |
| Rane et al. (2018) | + | + | + | + | + | - | + | + | + | 8/9 | B |
| Sundermann et al. (2018) | + | + | + | + | + | + | + | + | - | 8/9 | B |
| Dong et al (2019) | + | + | + | + | + | + | / | - | + | 7/9 | B |
| Lehtovirta et al. (1995) | + | + | - | + | + | + | + | / | - | 6/9 | B |
| Du et al. (2006) | + | / | + | + | + | + | + | / | + | 7/9 | B |
| Barber et al. (2000) | + | + | + | + | + | + | + | - | - | 7/9 | B |
| Boccardi et al. (2004) | + | + | + | + | + | + | + | / | - | 7/9 | B |
| Reiman et al. (1998) | + | + | + | + | + | + | + | / | - | 7/9 | B |
| Schmidt et al. (2008) | + | + | + | + | + | + | + | / | - | 7/9 | B |
| Basso et al. (2006) | + | + | + | + | + | + | + | + | / | 8/9 | B |
| Espeseth et al. (2006) | + | + | + | + | + | + | + | / | - | 7/9 | B |
| Tanaka et al. (1998) | + | + | - | + | + | + | + | / | - | 6/9 | B |
| Bigler et al. (2002) | + | + | + | + | + | - | + | + | - | 7/9 | B |
| Barber et al. (1999) | + | / | + | + | + | + | + | / | - | 6/9 | B |
| Carmelli et al. (2000) | + | / | + | + | + | + | + | / | + | 7/9 | B |
| Pennanen et al. (2006) | + | + | / | + | + | + | + | + | + | 8/9 | B |
| Morra et al. (2009) | + | + | / | + | + | + | / | + | + | 7/9 | B |
| Goltermann et al. (2019) | + | / | + | + | + | + | + | + | / | 7/9 | B |
| Konishi et al. (2018) | + | / | + | + | + | + | + | / | + | 7/9 | B |
| Kelly et al. (2018) | + | / | + | + | + | + | + | + | + | 8/9 | B |
| Alemany et al. (2018) | + | / | + | + | + | + | + | / | + | 7/9 | B |
| Tosun et al. (2010) | + | + | / | + | + | + | + | / | + | 7/9 | B |
| Hua et al. (2008) | + | + | / | + | + | + | + | + | + | 8/9 | B |
| Filippini et al. (2009) | + | + | / | + | + | + | + | / | + | 7/9 | B |
| Jak et al. (2007) | + | / | + | + | - | - | + | + | + | 6/9 | B |
| Doody et al. (2000) | + | / | - | + | + | + | + | / | - | 5/9 | B |
| Geroldi et al. (2000) | + | + | - | + | + | - | + | + | - | 6/9 | B |
| Mishra et al. (2018) | + | / | + | + | + | - | + | + | + | 7/9 | B |
| Bussy et al. (2019) | + | / | + | + | + | - | + | + | + | 7/9 | B |
| Taylor et al. (2014) | + | / | + | + | + | - | + | / | - | 5/9 | B |
| Kerchner et al. (2014) | + | + | + | + | + | - | + | + | + | 8/9 | B |
| Fennema-Notestine et al. (2011) | + | / | + | + | + | - | + | / | + | 6/9 | B |
| Andrawis et al. (2012) | + | + | / | + | + | + | + | + | + | 8/9 | B |
| Chang et al. (2016) | + | / | + | + | + | - | + | / | + | 6/9 | B |
| Honea et al. (2009) | + | / | + | + | + | + | + | / | + | 7/9 | B |
| Chang et al. (2014) | + | / | + | + | + | + | + | + | + | 8/9 | B |
| Li et al. (2016) | + | + | / | + | + | + | + | + | + | 8/9 |  |
| Okonkwo et al. (2010) | + | + | / | + | + | + | + | / | + | 7/9 | B |
| Tosun et al. (2011) | + | + | / | + | + | + | + | + | + | 8/9 | B |
| Tang et al. (2015) | + | + | / | + | + | + | + | + | + | 8/9 | B |
| Burggren et al. (2008) | + | / | + | + | - | - | + | / | - | 4/9 | B |
| Reiter et al. (2017) | + | / | + | + | + | + | + | + | + | 8/9 | B |
| Lampert et al. (2014) | + | / | + | + | + | + | + | + | + | 8/9 | B |
| Bender and Raz (2012) | + | / | + | + | + | + | + | / | + | 7/9 | B |
| Mueller and Weiner (2009) | + | + | - | + | + | + | + | / | - | 6/9 | B |
| Mueller et al. (2008) | + | + | / | + | + | + | + | + | + | 8/9 | B |
| Donix et al. (2010) | + | / | + | + | / | - | + | + | + | 6/9 | B |
| Ferencz et al. (2013) | + | / | + | + | + | + | + | / | - | 6/9 | B |
| Spampinato et al. (2011) | + | / | / | + | + | + | + | + | + | 7/9 | B |
| Ma et al. (2016) | + | / | + | + | + | + | + | / | + | 7/9 | B |
| Lu et al. (2011) | + | / | + | + | + | + | + | + | + | 8/9 | B |
| Dean et al. (2014) | + | / | + | + | + | / | + | / | + | 6/9 | B |
| Wolk et al. (2010) | + | + | / | + | + | + | + | / | + | 7/9 | B |
| Soldan et al. (2015) | + | / | + | + | + | + | + | + | - | 7/9 | B |
| Fan et al. (2010) | + | / | + | + | + | + | + | / | + | 7/9 | B |
| Rojas et al. (2018) | + | / | + | + | + | + | + | / | - | 6/9 | B |
| Banks et al. (2017) | + | / | + | + | + | + | + | / | + | 7/9 | B |
| Walsh et al. (2013) | + | / | + | + | + | + | + | / | - | 6/9 | B |
| Li et al. (2017) | + | + | / | + | + | + | + | + | + | 8/9 | B |
| Chen et al. (2012) | + | / | + | + | + | + | + | / | + | 7/9 | B |
| Hostage et al. (2014) | + | + | - | + | + | / | + | / | + | 6/9 | B |
| Donix et al. (2013) | + | + | / | + | + | / | + | + | - | 6/9 | B |
| Shi et al. (2014) | + | + | / | + | / | / | + | / | + | 5/9 | B |
| Hafsteinsdottir et al. 2012 | + | + | / | + | + | - | + | + | - | 6/9 | B |
| Novellino et al. (2019) | + | + | - | + | - | - | + | + | + | 6/9 | B |
| Juottonen et al. (1998) | + | + | / | + | - | - | / | + | - | 4/9 | B |
| Wilhelm et al. (2008) | + | - | - | - | + | + | / | / | - | 3/9 | B |
| Cherbuin et al. (2008) | + | + | - | - | + | + | + | - | + | 6/9 | B |
| Biffi et al. (2010) | + | + | / | + | + | + | + | + | + | 8/9 | B |
| Ystad et al. (2009) | + | / | / | + | + | - | / | / | + | 4/9 | B |
| Schuff et al. (2009) | + | + | + | + | + | - | / | + | + | 7/9 | B |
| Stewart et al. (2011) | + | + | + | + | + | + | / | / | / | 6/9 | B |
| Protas et al. (2013) | + | / | - | - | + | + | + | / | / | 4/9 | B |
| Hoogendam et al. (2012) | + | + | + | + | + | + | + | / | + | 8/9 | B |
| Geroldi et al. (1999) | + | + | - | + | + | - | / | + | / | 5/9 | B |
| Lehtovirta et al. (1996) | + | + | / | + | + | + | / | / | / | 5/9 | B |
| O’Dwyer et al. (2012) | + | + | / | + | + | + | / | / | + | 6/9 | B |
| Sabuncu et al. (2012) | + | + | / | + | + | + | / | + | + | 7/9 | B |
| Bunce et al. (2012) | + | + | + | + | + | - | + | / | + | 7/9 | B |
| Hostage et al. (2013) | + | + | / | + | + | - | / | / | + | 5/9 | B |
| DiBattista et al. (2014) | + | + | / | + | / | / | - | - | - | 3/9 | B |
| Manning et al. (2014) | + | + | - | - | + | + | / | / | + | 5/9 | B |
| Khan et al. (2014) | + | - | - | + | - | - | - | - | + | 3/9 | B |
| Holland et al. (2013) | + | + | - | - | + | + | / | / | + | 5/9 | B |
| Lyall et al. (2013) | + | + | + | + | + | + | + | / | + | 8/9 | B |
| Morgen et al. (2015) | + | + | + | + | - | - | + | + | + | 7/9 | B |
| Risacher et al. (2015) | + | + | - | + | + | + | / | / | - | 5/9 | B |
| Yokoyama et al. (2015) | + | + | + | + | + | + | + | / | - | 7/9 | B |
| Sampedro et al. (2015) | + | / | - | + | + | + | - | / | - | 4/9 | B |
| Khan et al. (2017) | + | + | - | + | + | + | - | / | + | 6/9 | B |
| Falahati et al. (2017) | + | / | - | + | + | - | - | + | + | 5/9 | B |
| Rogne et al. (2016) | + | + | + | + | + | + | / | + | - | 7/9 | B |
| Konishi et al. (2016) | + | - | - | + | + | + | + | / | - | 5/9 | B |
| Habes et al. (2016) | + | + | + | + | + | + | + | + | + | 9/9 | B |
| Fang et al. (2019) | + | + | - | + | + | - | + | + | - | 6/9 | B |
| Lupton et al. (2016) | + | + | - | + | + | + | - | - | + | 6/9 | B |
| Hobel et al. (2019) | + | / | + | + | + | + | + | / | + | 7/9 | B |
| Schreiber et al. (2017) | + | + | - | + | + | + | - | - | + | 6/9 | B |
| Haller et al. (2017) | + | + | + | + | + | + | + | + | + | 9/9 | B |
| Nao et al. (2017) | + | / | + | + | + | + | + | / | - | 6/9 | B |
| Li et al. (2019) | + | + | - | + | + | - | + | + | - | 6/9 | B |
| Hays et al. (2019) | + | + | + | + | + | + | + | / | + | 8/9 | B |
| Herrmann et al. (2019) | + | + | + | + | + | + | + | / | - | 7/9 | B |
| Foley et al. (2017) | + | / | + | + | - | - | + | / | + | 5/9 | B |
| Ghisays et al. (2019) | + | + | + | + | + | + | + | / | - | 7/9 | B |
| Cotta Ramusino et al. (2019) | + | / | / | / | + | + | + | / | + | 5/9 | B |
| Lyall et al. (2019) | + | / | / | / | + | + | + | + | + | 6/9 | B |

LOE: level of evidence; -: score not fulfilled; +: score fulfilled; /: answer is not clear

NOS Scale: (Items 1 to 4 classified under Selection category). 1 = Is the case definition adequate?; 2 = Representativeness of the cases; 3 = Selection of controls; 4 = Definition of controls; (Items 5 and 6 under Comparability category) 5 = Study controls for age or sex; 6 = Study controls for any additional factor; (Items 7, 8, and modified 9 under Exposure category) 7 = Ascertainment of exposure; 8 = Same method of ascertainment for cases and controls; 9 = Visual inspection of the MRI data quality.

Table 4 LOE, according to the 2005 classification system of the Dutch Institute for Healthcare Improvement CBO ((www.cbo.nl)

|  | Intervention |
| --- | --- |
| A1 | Systematic review of at least 2 independent from each other conducted studies of evidence level A2 |
| A2 | Randomized double-blinded comparative clinical research of good quality and efficient size |
| B | Comparative research, but not with al characteristics as mentioned for A2. This includes also patient-control research and cohort research. |
| C | Not comparative research |
| D | Opinion of experts |

Table 5 Strength of Conclusion (modified table)

|  | Conclusion based on |
| --- | --- |
| 1 | Research of evidence level A1 or at least 2 independent conducted studies of evidence level A2 |
| 2 | 1 research of evidence level A2 or at least 2 independent conducted studies of evidence level B |
| 3 | 1 research of evidence level B or C |
| 4 | Opinion of experts or Inconclusive or inconsistent results between various studies |
