## Supplementary material for "Apolipoprotein E genotype and MRI-detected brain alterations pertaining to neurodegeneration: A systematic review": Figure 1

Figure 1. PRISMA flowchart of the study selection process

Additional records identified through other sources

(n = 17)

Records excluded

(n = 236)

Studies included in qualitative synthesis

(n = 115)

Full-text articles assessed for eligibility

(n = 239)

Records identified through database searching: Pubmed (n = 693); Ovid (n = 48); Scopus (n = 42); Cochrane (n = 0)

Records screened on titles and abstracts

(n = 475)

Records after duplicates removed

(n = 475)

Full-texts articles excluded (n = 124)

Reasons:

- Populations (n = 72)
- Outcome (n = 26)
- Design (n = 26)
